# Digital Health Technologies for Alzheimer’s Disease and Related Dementias: Initial Results from a Landscape Analysis and Community Collaborative Effort

**DOI:** 10.1101/2024.03.18.24304471

**Authors:** Sarah Averill Lott, Emmanuel Streel, Shelby L. Bachman, Kai Bode, John Dyer, Cheryl Fitzer-Attas, Jennifer C. Goldsack, Ann Hake, Ali Jannati, Ricardo Sainz Fuertes, Piper Fromy

**Author notes:** Corresponding author: Sarah Averill Lott.

## Abstract

Digital health technologies offer valuable advantages to dementia researchers and clinicians as screening tools, diagnostic aids, and monitoring instruments. To support the use and advancement of these resources, a comprehensive overview of the current technological landscape is essential.

A multi-stakeholder working group, convened by the Digital Medicine Society (DiMe), conducted a landscape review to identify digital health technologies for Alzheimer’s disease and related dementia populations. We searched studies indexed in PubMed, Embase, and APA PsycInfo to identify manuscripts published between May 2003 to May 2023 reporting analytical validation, clinical validation, or usability/feasibility results for relevant digital health technologies. Additional technologies were identified through community outreach. We collated peer-reviewed manuscripts, poster presentations, or regulatory documents for 106 different technologies for Alzheimer’s disease and related dementia assessment covering diverse populations such as Lewy Body, vascular dementias, frontotemporal dementias, and all severities of Alzheimer’s disease. Wearable sensors represent 32% of included technologies, non-wearables 61%, and technologies with components of both account for the remaining 7%. Neurocognition is the most prevalent concept of interest, followed by physical activity and sleep. Clinical validation is reported in 69% of evidence, analytical validation in 34%, and usability/feasibility in 20% (not mutually exclusive).

These findings provide clinicians and researchers a landscape overview describing the range of technologies for assessing Alzheimer’s disease and related dementias. A living library of technologies is presented for the clinical and research communities which will keep findings up-to-date as the field develops.

## Introduction

The uses for digital health technologies (DHTs) for clinical research, personalized medicine, and general wellness are well established and continue to expand [1, 2]. In the field of Alzheimer’s disease and related dementias (ADRD), growth in the development and use of DHTs [3] is occurring alongside increased research and development of drugs for the prevention and treatment of Alzheimer’s disease (AD) and other dementia-inducing disorders including Lewy Body and frontotemporal dementias[4–6]. The recent regulatory approval of some therapeutic agents targeting AD pathophysiology highlights the complexity of dementia management, as new treatment opportunities emerge alongside ongoing challenges in dementia research, clinical management, and holistic patient care [7]. These challenges include identifying digital biomarkers that signal changes early in the disease course when intervention is more likely to provide a substantial impact; assessing clinical manifestations of dementia such as activities of daily living (ADLs), cognitive function, physical activity, and sleep; evaluating and demonstrating response to therapeutic interventions [8]; and improving recruitment (including recruitment of diverse patient groups [9], retention, and assessment of participants in clinical trials). In addressing these challenges, clinicians and researchers will benefit from utilizing DHTs as fit for purpose tools that can play a strategic role in screening, monitoring, and longitudinal assessment in research, clinical, and real world settings.

Members of a global, multi-stakeholder working group convened by the Digital Medicine Society (DiMe)[10] met over several months to assess the characteristics of DHTs in ADRD populations, recognizing the immense opportunity for further research and clinical impact alongside the ever-increasing burden of Alzheimer’s disease and related dementias[11, 12]. DiMe, a global non-profit dedicated to driving scientific progress and broad acceptance in digital medicine to redefine healthcare and improve lives, hosts the Digital Health Measurement Collaborative Community (DATAcc)[13], a U.S. Food and Drug Administration (FDA) Center for Diagnostic and Radiological Health (CDRH) collaborative community[14], and convenes a wide range of stakeholders and subject matter experts in pre-competitive forums to advance the field.

While others have examined the evidence for ADRD-relevant DHTs[15, 16], such efforts are often limited to a subset of available technologies or therapeutic domains, obscuring substantial portions of the DHT landscape and restricting opportunities for translation of the benefits of these tools across different dementias. To obtain a more comprehensive view, our ADRD working group aggregated multiple technologies across ADRD-therapeutic areas by surveying 20 years of peer-reviewed literature and then sourcing additional evidence from DHT users and developers. While we were primarily informed by the literature and had strict inclusion and exclusion criteria, our purpose in this exercise was to identify and collect digital health technologies in use for dementia measurement, not to interrogate the literature for quality or bias as one would expect in a traditional systematic literature review. Rather, our broad landscape assessment was literature-based and community supported, as the DiMe working group (comprised of ADRD experts from clinical, pharmaceutical, industry research and development, and patient advocacy domains; see acknowledgments for complete list of participating organizations) were given an opportunity to review the findings and identify additional technologies for consideration.

The results of this innovative effort are reported here, bringing an open science perspective and outlining the state of digital innovation in ADRD research. As a precompetitive group representing a range of interests, we do not promote any specific technology type, product, or developer in this analysis and therefore refrain from providing direct product examples in-text; a list of all sources that inform this overview is provided in the supplement so that interested readers may seek specific examples if desired. Here we provide a general overview advancing awareness of the range of technologies and their uses for dementia assessment.

## Methods

### Data Sources

The working group sought to identify DHTs with associated evidence of validity[17] and therefore started by turning to the peer-reviewed literature. In May 2023, we conducted a landscape review to identify DHTs being used for assessment in ADRD populations by the clinical and research communities. The search strings (*see supplementary material*) included terminology to identify DHTs, diagnostic and descriptive terms to focus on ADRD-relevant populations, and pertinent concepts of interest measured by a DHT including sleep health, memory, language, social function, oculomotor function, cognition, life space mobility, essential bodily functioning, and emotional or behavioral concerns. The selection of these concepts of interest was informed by literature focused on meaningful aspects of health[8, 18, 19] for ADRD stakeholders at large. We restricted search results to primary research studies with human participants published within the last 20 years. To avoid over-representation of any given technology brand, we intentionally did not include any name-brand affiliated keywords in the search terms, knowing this might mean relevant studies were omitted from the results if they included only brand identifiers for utilized technologies. The search string was performed within PubMed, APA PsycInfo, and Embase literature databases, and the results were then aggregated and deduplicated within reference management software Zotero[20].

### Literature Review Screening

Identified articles were screened by two reviewers (SAL, PF), with a random sample of 5% co-reviewed to ensure harmony on inclusion/exclusion standards. Articles were included or excluded based on the criteria outlined in Table 1.

**Table 1.**
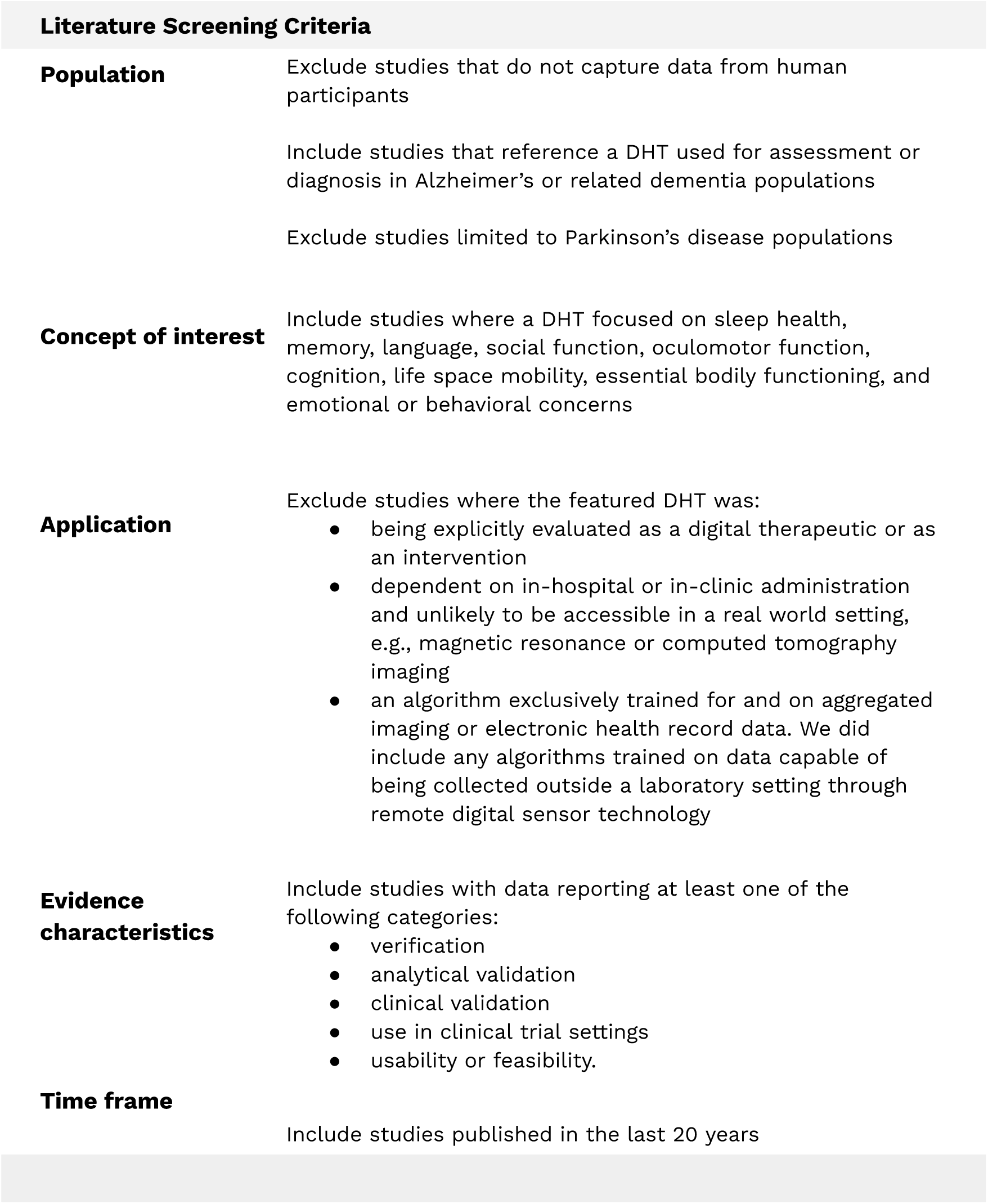
Inclusion/exclusion criteria applied to papers identified in literature search or returned by survey.

### Extraction

From each qualifying manuscript, we extracted the name and manufacturer of the technology being utilized; the technology type, form factor, and wear location; the reported therapeutic category and population descriptors as described verbatim by study authors; and the general health concepts and outcomes under assessment. Categories used elsewhere in assessing and categorizing DHTs informed our classification labels[21, 22] for technology types and health outcomes. Technology types were sorted into ambient (standalone, not worn on the body), wearable (worn on the body for any period of time), implantable (surgically placed), or ingestible (swallowed); while form factor descriptions were as provided by study authors in the text. Broad health outcome categories were sorted into activities of daily living, mental health, neurocognitive, neurological or sensory, physical activity, and sleep.

More specific health outcome category labels were derived from study descriptors and are also reported. Lastly, we labeled evidence according to the type of data it reported: verification, analytical validation, clinical validation, and usability or feasibility[17]. When extracting these evidence types and other characteristics such as diagnostic classifications, study populations descriptors, or outcome measures, we adhered to author/publication verbiage as closely as possible with the goal of collating rather than interpreting labels or assessing individual pieces of evidence as to their rigor or performance.

### Community Sourcing

ADRD subject matter experts in DHT development, clinical trials, and clinical care from the DiMe ADRD project team[10] reviewed and added to the list of technologies created during extraction, to identify additional technologies not captured in our literature search. Developers of identified DHTs were contacted by email (when contact information was accessible) and provided a survey via a Qualtrics[23] link where they could return peer reviewed articles, conference posters or presentations, regulatory documents, datasets, open-source algorithms, or other supporting materials for their technologies in line with the V3 framework [17]. If a developer responded and provided any evidence, that data was assessed against the same inclusion and exclusion standards for the literature presented in Table 1 and included in our results if eligible. Finally, the survey link and a description of the library were shared on LinkedIn during World Alzheimer’s Month in an effort to solicit evidence for ADRD DHTs from the broader scientific and digital health communities (September, 2023).

## Results

The combined literature search resulted in an initial dataset of 1,743 manuscripts; after screening we identified 153 eligible articles related to 117 unique DHTs. We reviewed and omitted 13 technologies that were clearly obsolete and discontinued, bringing the results from the literature to 102 DHTs. Project partners reviewed the complete list and made us aware of an additional 57 technologies, resulting in a potential total of 159 unique DHTs for ADRD assessment. Surveys were disseminated via email to technology developers of the additional technologies to request evidence so their technologies could be considered for inclusion; as of this writing, 42 survey submissions featuring 17 technologies were returned. Survey responses included peer-reviewed publications, conference poster and oral presentation materials, and documentation of regulatory approvals. After assessing all returned materials for eligibility as outlined in Table 1, our total results included 172 pieces of evidence representing 106 unique DHTs. A list of the results of eligible manuscripts/evidence and the respective technologies therein is available in the supplementary material.

### Populations Represented

Our results captured a wide variety of ADRD population descriptors as reported by study authors or DHT developers (Table 2). The classifications most represented include Mild Cognitive Impairment (MCI) in 39% of evidence records and 49% of technologies, followed by the general classification of Alzheimer’s disease indicated in 31% of evidence records and 40% of technologies.

**Table 2.**
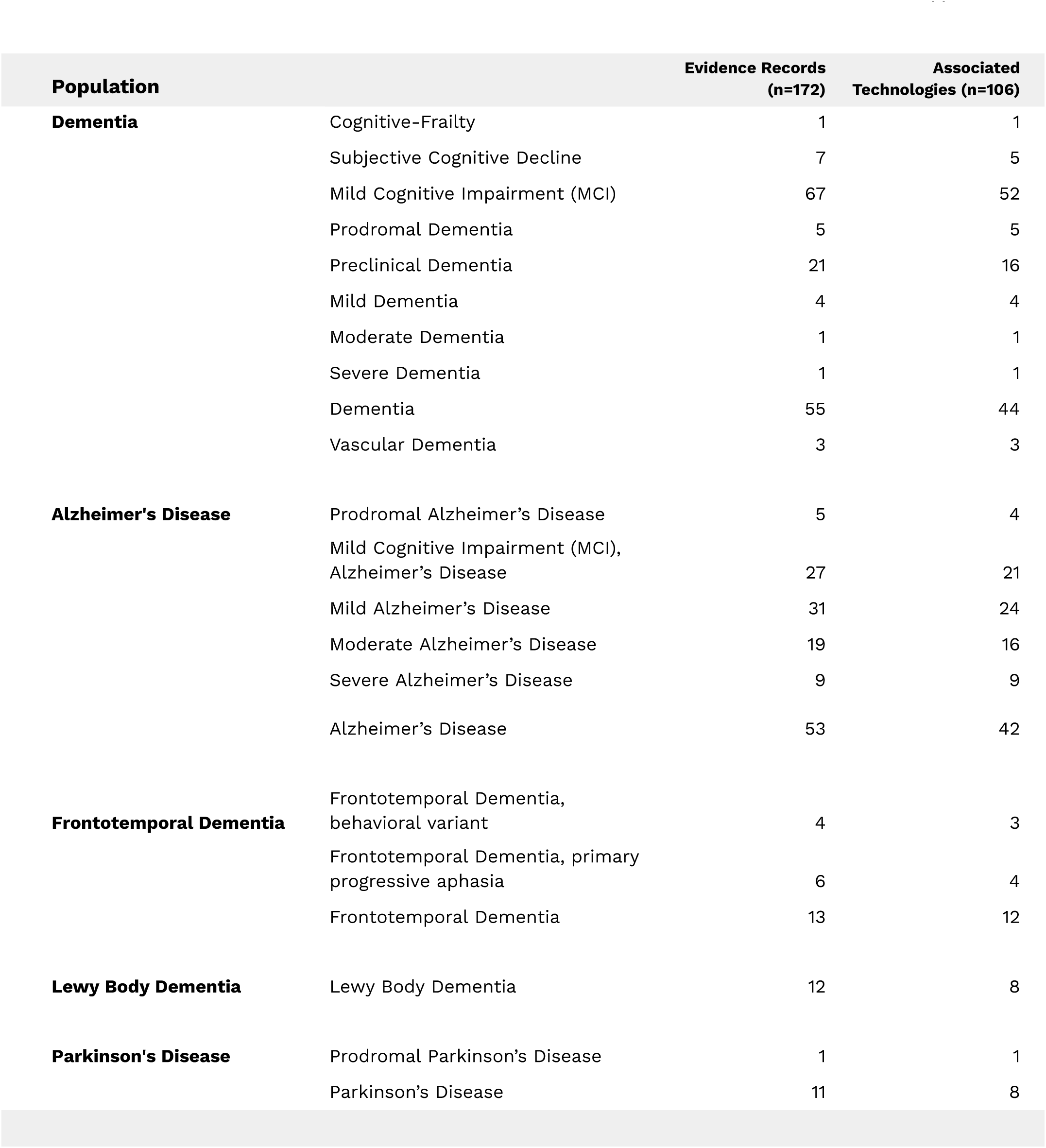
Frequencies of population areas of focus in the library, as reported in manuscripts. Many pieces of evidence reference multiple diagnostic classifications.

### Technology Types, Form Factors, and Health Concepts

Ambient (or non-wearable sensor technologies) are the most prevalent technology type, representing 61% of included DHTs and 68% of the evidence (Table 3). This category includes environmental or stand alone sensors as well as software and application-based technologies which are not tied to a given device. 83% of ambient technologies fall into the software and application category. Wearable sensor devices represent 32% of DHTs and 26% of evidence. Seven percent of technologies incorporate both wearable and ambient technologies. The most common form factors are software and applications designed for a tablet or smartphone (42% DHTs; 45% of evidence) and sensors worn on the body but not as a smartwatch (e.g. attached via a strap or brace) (19% DHTs; 16% of evidence). Cameras and contactless sensors are also strongly represented in the ambient category (collectively 13% DHTs; 14% evidence), while smartwatches represent a smaller portion of the results (8% DHTs; 5% evidence). Most studies feature commercially available DHTs (89% of evidence) though we included experimental/noncommercial DHTs if relevant to our population and concepts of interest (11% evidence). Our results also returned examples of “smart home” applications with a combination of different technology types designed to passively monitor life space mobility such as cameras, movement detecting radar, or pressure mats.

**Table 3.**
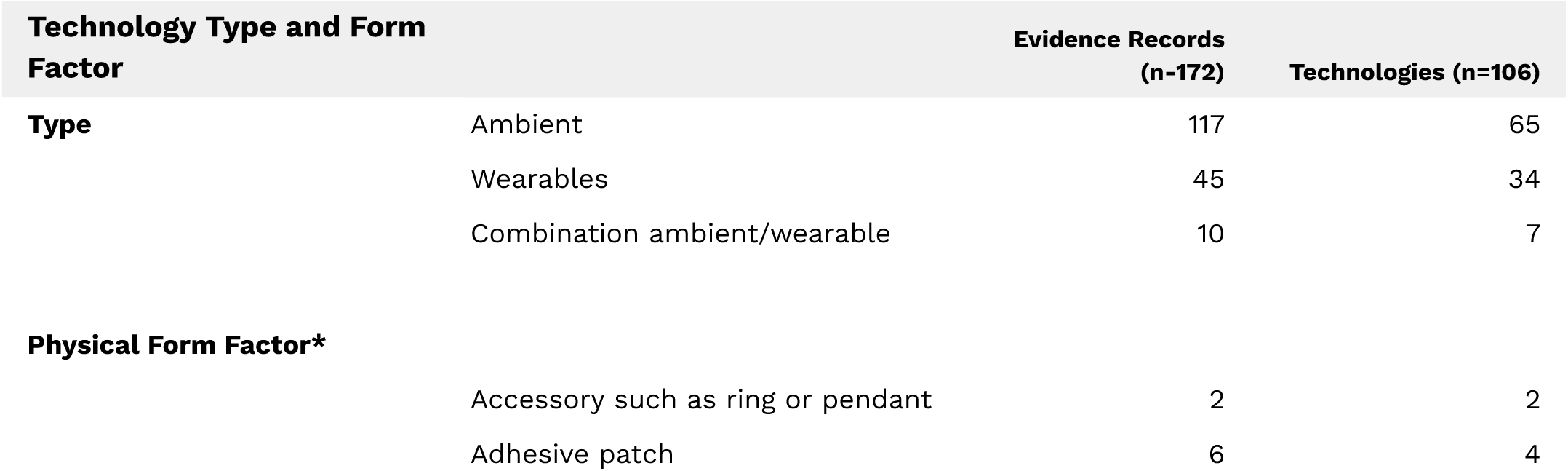
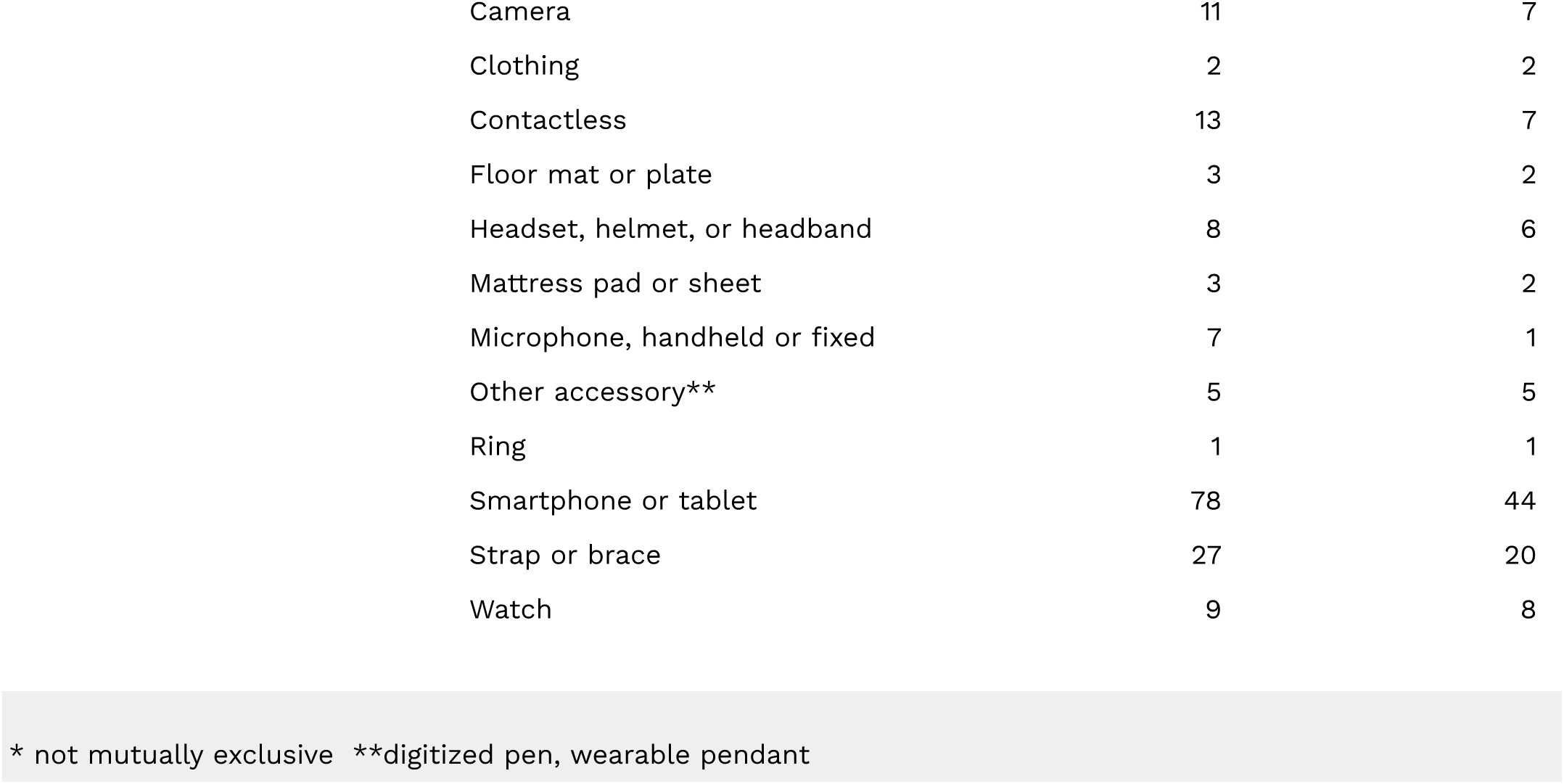
Prevalent technology types and form factors represented in the ADRD DHT library. Ambient includes environmental and software or application-based technologies.

DHTs are classified into the health concepts of interest they assess, including but not limited to activities of daily living; mental health, neurocognitive function, neurological or sensory function, physical activity, and sleep (Table 4). The most frequent concept of interest is neurocognitive function with 43% of technologies and 51% of the evidence, but physical activity represents a larger portion of DHTs (48%) and is addressed in 42% of evidence. Measured outcomes as reported in included studies were analyzed to categorize the evidence into more specific health concepts. Memory is the concept of interest addressed most often as a primary focus of 26% of DHTs and 26% of evidence, but life space mobility, speech patterns and characteristics, gait and mobility, sleep, and several subdomains of memory and cognitive function are also well represented (Table 4).

**Table 4.**
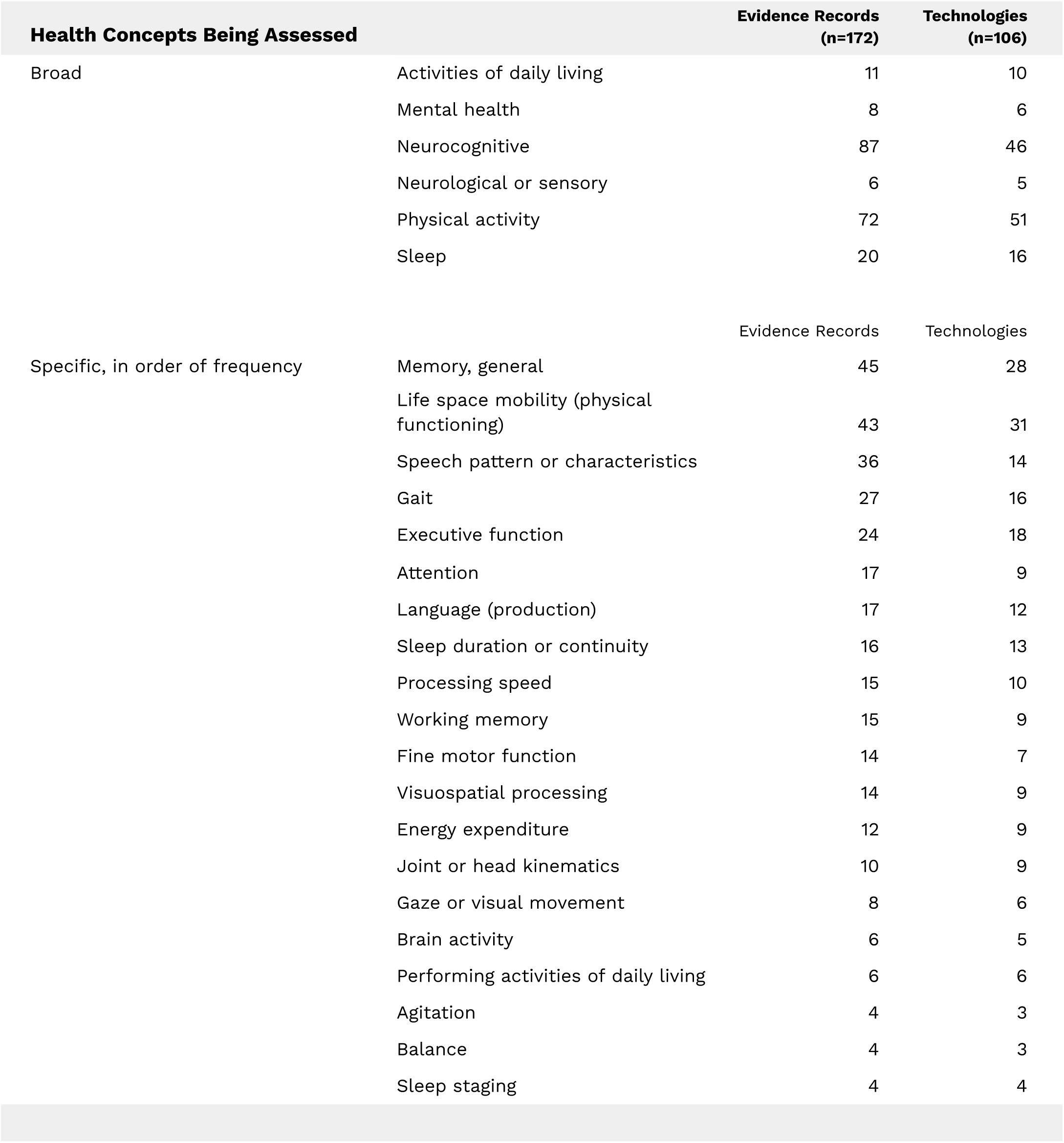
Prevalent concepts of interest in the ADRD DHT library. These are not mutually exclusive. These figures represent frequency in library evidence along with the number of corresponding technologies as of October 2023.

### V3, other evidence standards in action

The V3 framework[17] codifies best practices for evaluating verification, analytical validation, and clinical validation to determine whether DHTs are fit for purpose. Briefly, these processes evaluate whether sensor-based technologies and their algorithm(s) measure and interpret what they intend to measure, and provide metrics that are clinically or functionally meaningful in the stated context of use[17]. To reflect these standards, we categorized evidence to indicate when it reports verification, analytical validation, clinical validation, and/or usability. Most of the current evidence reports validation: 34% of the evidence reports analytical validation while a majority (69%) reports evidence for clinical validation within ADRD populations. 19% percent of the evidence reports demonstrated usability or feasibility within ADRD populations. Just three manuscripts reported verification. These categories are not mutually exclusive; a single manuscript might report on multiple aspects.

## Discussion

To the best of our knowledge, this review is the first to broadly document the growing landscape of DHTs available for assessment within the ADRD space. DHTs focused on cognition and memory are unsurprisingly prevalent in our results. However, we identified several other concepts of interest in the literature, underscoring the versatility DHTs offer for holistic assessment of ADRD populations. When examining technology types, mobile or software applications are prevalent and their broad scope is notable. We find these technologies are being used by researchers and clinicians to assess multiple aspects of neurocognition (including several types of memory, attention, executive functioning, and processing speed); speech production and language characteristics; fine motor functioning; oculomotor behavior; sleep; gait, mobility, or balance; life space mobility dynamics; and emotional or other behavioral patterns. Given the ubiquity of smartphones and web-based options for assessment, these findings indicate a large capacity for sensitive and ongoing patient assessment or monitoring in clinical and real world settings, including the critical opportunity to reach patients not previously accessible to research or therapeutic settings. A majority (89%) of our results feature commercially available DHTs, emphasizing that a range of technological solutions are available and in use by clinicians and researchers even as additional development and innovation continues.

### Biomarkers, early detection opportunities, and digital phenotyping

Our results indicate researchers are leveraging DHTs to identify digital biomarkers and behavioral changes that can provide early indicators of functional and cognitive changes in both clinical and remote settings. Examples include but are not limited to wearable headbands for in-home EEG assessment to monitor sleep, sophisticated analysis of speech patterns and vocal dynamics as captured by smartphone speakers, cameras tracking patterns in narrow applications (gaze/eye movement) as well as broad (behavioral) patterns in the home; and inertial sensors worn on the lower back, chest, arm, or leg to monitor mobility patterns. These technologies detect subtle functional and behavioral differences that can differentiate patients from healthy controls and discriminate between disease stages. When tablet or smartphone apps, wearables, and other DHTs are used in aggregate, the results can be quite illustrative, providing a “digital phenotype” [24] that captures a comprehensive in-situ portrait of someone’s daily functional and cognitive status, providing a better window into a patient’s emotional state, social patterns, and physical experiences to generate research insights and guide clinical care in ways that are meaningful to patients[25]. “Smart home” applications show promise in this regard as well, with personalized and multimodal monitoring that might include cameras that track life space mobility paired with bed pressure mats to detect sleep patterns, restlessness, or agitation.

When these technologies detect signals such as behavioral and activity changes, a digital phenotype can be established. Such setups are currently utilized in a variety of ways including discriminating between disease states[26], detecting agitation or apathy[27], unobtrusively monitoring response to therapeutics in care settings[28], and improving the selection of candidates for clinical research purposes[29].

### Clinical research

The importance for understanding the landscape of ADRD-based DHTs is apparent when considering clinical research. Broadly, DHTs can improve trials by (a) reducing within-participant measurement noise [30], ultimately leading to improved statistical power, smaller trials[29], and faster paths to regulatory judgment of new therapies and (b) decentralizing assessments, enabling the participation of individuals located in more diverse geographical areas and therefore acquiring a more representative and diverse sample of the population under study[9]. (For explication of this approach, see Sliwinski, 2008)[31]. Although there is a recent example elsewhere in clinical research using a DHT as a primary clinical endpoint[32], this is not current practice in the ADRD space. However, the growing acceptance of DHTs as secondary endpoints in ADRD research[33, 34] is slowly moving the field towards this possibility[35, 36]. Studies employ DHTs for monitoring response to treatment in a variety of ways, such as using actigraphy to discern impacts of Mevidalen on activity and sleep within Lewy Body dementia cohorts[37, 38] or using smartwatches to detect suvorexant-induced changes in sleeping patterns for persons with Alzheimer’s experiencing insomnia[39].

### Digital neuropsychology

The dominance of technologies assessing memory (short term, long term, working), executive function, attention, processing speed, verbal fluency, and visuospatial processing document an increasingly digital shift within neuropsychological assessment. Clinicians see value in the opportunity to assess functional status in real world settings[40] even as they grapple with concerns around ecological validity[25, 41] due to variations in technology, computer literacy, environmental factors, and the presence or lack of supervision by a trained observer[41, 42]. While further research is needed to assuage these concerns, data continues to emerge illustrating that innovative digital assessment modalities can successfully discriminate between healthy controls and individuals in early stages of Alzheimer’s disease[43]. Such DHTs provide clear advantages with regard to scalability and increasing access to patients for whom an in-person assessment may not be feasible[41], as well as the potential for real world and longitudinal evidence gathering[44] in populations where early detection is key. Many of the ambient technologies captured in this analysis were smartphone or tablet applications designed to analyze and interpret cognition or fine motor function to detect cognitive decline or discriminate between dementia states. Some examples include digital tasks testing aspects of cognition such as working memory, processing speed, or episodic memory[45]; executive function[46]; reaction time [47]; speech or text patterns[48, 49]; or pressure of a digital pen[50] on a device.

### Wearables versus ambient sensors

Wearable sensors generally (32%), and smartwatches in particular (8%) were less represented in our landscape review than ambient technologies. Given their abundance and relative accessibility, wearable sensors can play an important role in the sensitive assessment of many ADRD-relevant measures, including mobility[51] (including but not limited to gait, life space pattern changes, and fall detection) and other biomarkers such as late-onset essential tremor[52] or heart rate variability[3]. However, the lower proportion of wearables to ambient technologies may be a reflection of the versatility and advantages of the latter for dementia populations. For example, ambient sensors may be more appropriate for populations where memory or awareness concerns might make adherence with wearables a challenge. The aforementioned smart home possibilities with cameras, pressure sensors, and/or smart speakers/smartphones that detect changes in behavioral patterns or vocal biomarkers offer a powerful modality for real world continuous data collection while posing minimal burden on patients or caregivers.

### Diagnostic classifications

Our results encompass a wide spectrum of dementia diagnoses and severity classifications. However, we note the prevalence of Alzheimer’s disease in our results compared to Frontotemporal dementias and Lewy Body dementia, as well as the prevalence of diagnoses like mild cognitive Impairment or general dementia as compared to results that specifically examined populations with moderate or advanced diagnoses. This suggests an opportunity for DHT developers and researchers to study additional uses and clinical validity within populations that are more narrowly defined within disease severity class, or in dementias other than Alzheimer’s disease.

### Digital Measurement Products Library: A living resource

The digital health technology landscape evolves rapidly. Technologies from this review with peer-reviewed evidence published 2020-present are collated into the Library of Digital Measurement Products, an open access resource hosted by DiMe[53] for the clinical, research, and developer communities. While our landscape analysis included evidence published between 2003-2023 featuring DHTs that are both commercial and experimental, the library is a living resource intended to assist researchers and clinicians who may be considering a current and specific use case. Thus, it describes DHTs that are commercially available on the market at the time of entry into the database, and is restricted to peer-reviewed evidence published from 2020-present. Evidence is tagged according to the study type by which it is reported in the published literature as verification, analytical validation, clinical validation, or usability[17]. Categorical fields are filterable so the end user can sort and view evidence according to a specific technology or technology type, therapeutic area or concept of interest, or by reported evidence type. This library aligns with the mission of the Digital Health Measurement Collaborative Community[13, 14] to share knowledge and resources to advance digital medicine and ultimately improve patient lives; we encourage the wider ADRD community to join our collective efforts to ensure diverse evidence-supported DHTs are represented in this important resource. A link to submit evidence for potential inclusion into the library is available to the public on DiMe’s Library of Digital Measures Products web page[53]; beginning mid-2024, the library will be updated quarterly via review of community submissions, revisiting of the published literature, and removal of outdated evidence.

### Limitations

While our results provide a broad overview of DHTs for ADRD assessment across 20 years of literature, we intentionally excluded brand-name identifiers from our search strings to avoid bias by disproportionately representing some market technologies over others. Additionally, we omitted search terms specific to Parkinson’s disease dementia because it has differentiating features from other dementias, and therefore will be the focus of a similar exercise in future research. Due to this intentional limiting of some search terms, we do not claim the search results to be exhaustive. As we conducted this exercise to aggregate DHTs and their uses within ADRD, with a focus on extracting technologies rather than conducting a formal appraisal of the literature, we report these findings as a technology landscape analysis rather than a traditional systematic review.

Lastly, members of the DiMe working group made the authorship team aware of additional technologies which were not identified/discovered via the literature search.

While this carries a risk of bias (primarily that some additional technologies were presumably overlooked due to being unknown to project partners), we considered that the benefits of greater inclusion outweighed this potential drawback. We reached out to solicit supporting evidence for these, but received evidence back for a small portion. Consequently, many of the technologies identified during partner review are not included in this analysis, as we lacked the associated evidence necessary to report on them.

## Conclusion

DHTs show great promise for ADRD populations by enabling objective, continuous, and repeatable measurements of functioning and symptoms in a range of settings. Ambient, application-based, and wearable technologies offer vast real world and longitudinal evidence generation possibilities, as well as the opportunity to take traditional neuropsychological and neurocognitive assessment out of the clinic or laboratory and into homes and care settings. They offer additional information to facilitate clinical decision making, and provide opportunities for clinical research to identify earlier signals of disease onset, monitor disease trajectory, or detect response to therapeutic treatment at a time when demand is increasing for precision medicine that better identifies and targets appropriate candidates for care and intervention, particularly in the prodromal or earliest stages[54, 55]. These technologies continue to elucidate insights about domains such as cognitive function, sleep, mobility, language changes, and social and behavioral trends which will prove invaluable alongside traditional dementia research and management. This review confirms the multifaceted uses for and significant potential of DHTs as clinicians and researchers seek clarity and insights beyond the reach of traditional assessment modalities.

## Data Availability

All data produced in the present study are available upon reasonable request to the authors.

## Supplementary Material: Search terms; technologies and citations identified

a. Search terms

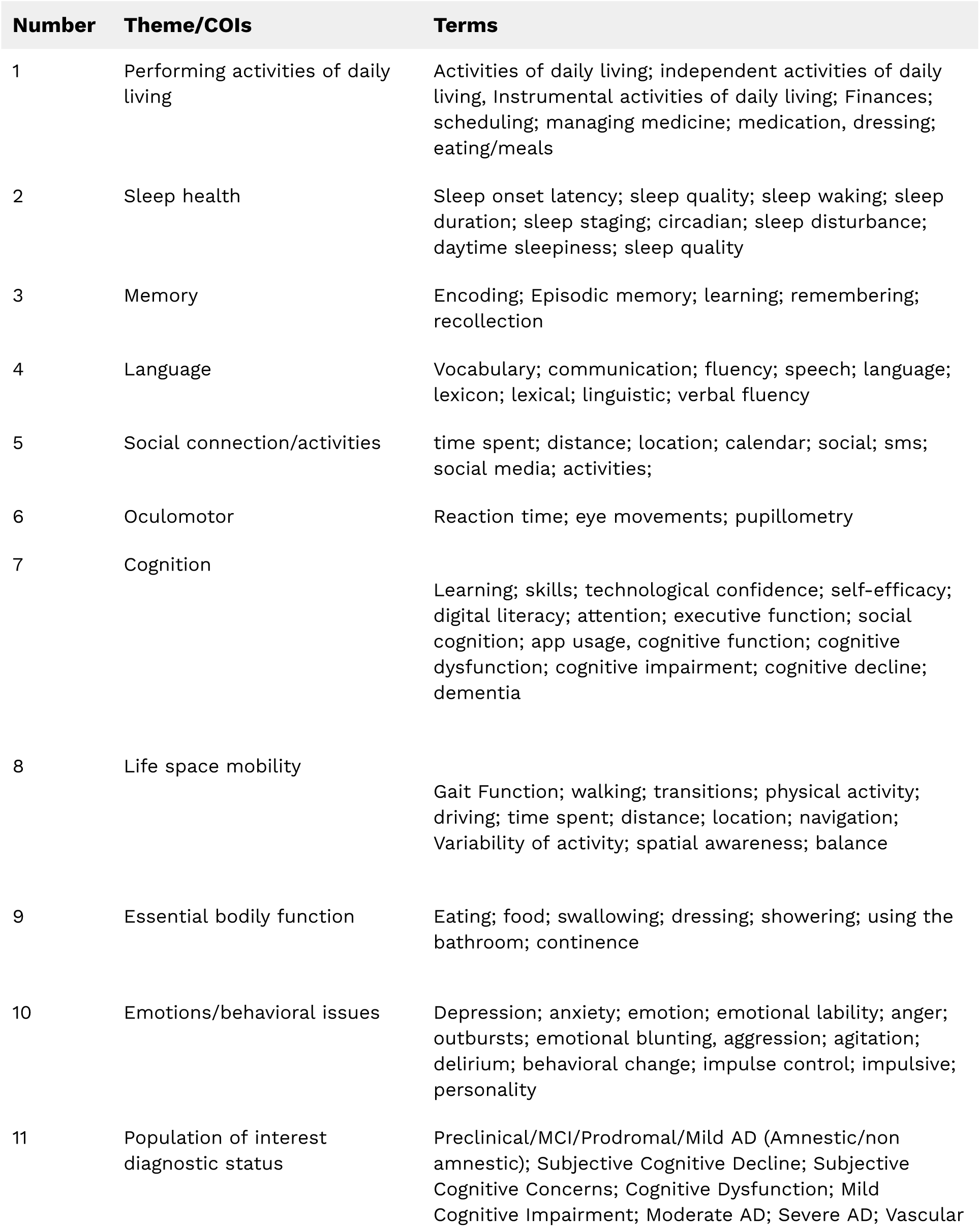
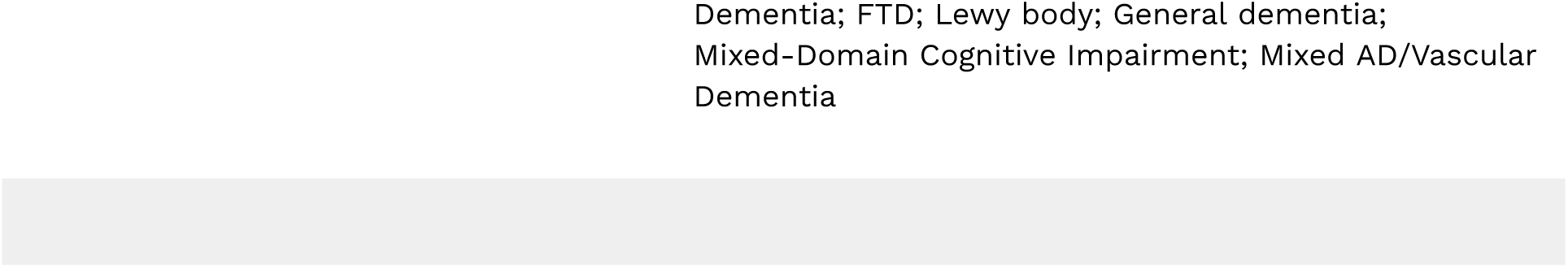

b. Technologies and evidence captured through literature search & partner review

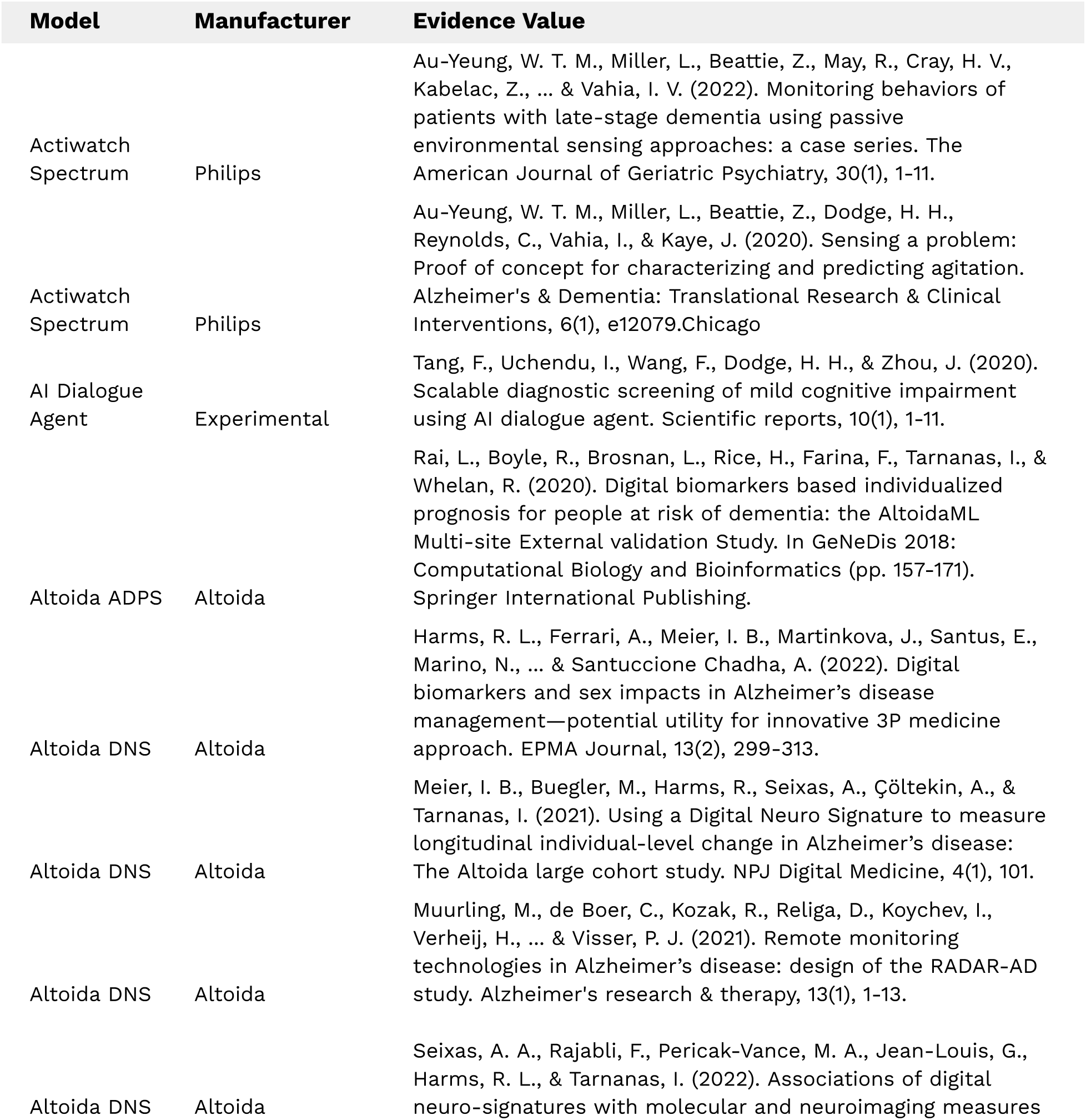
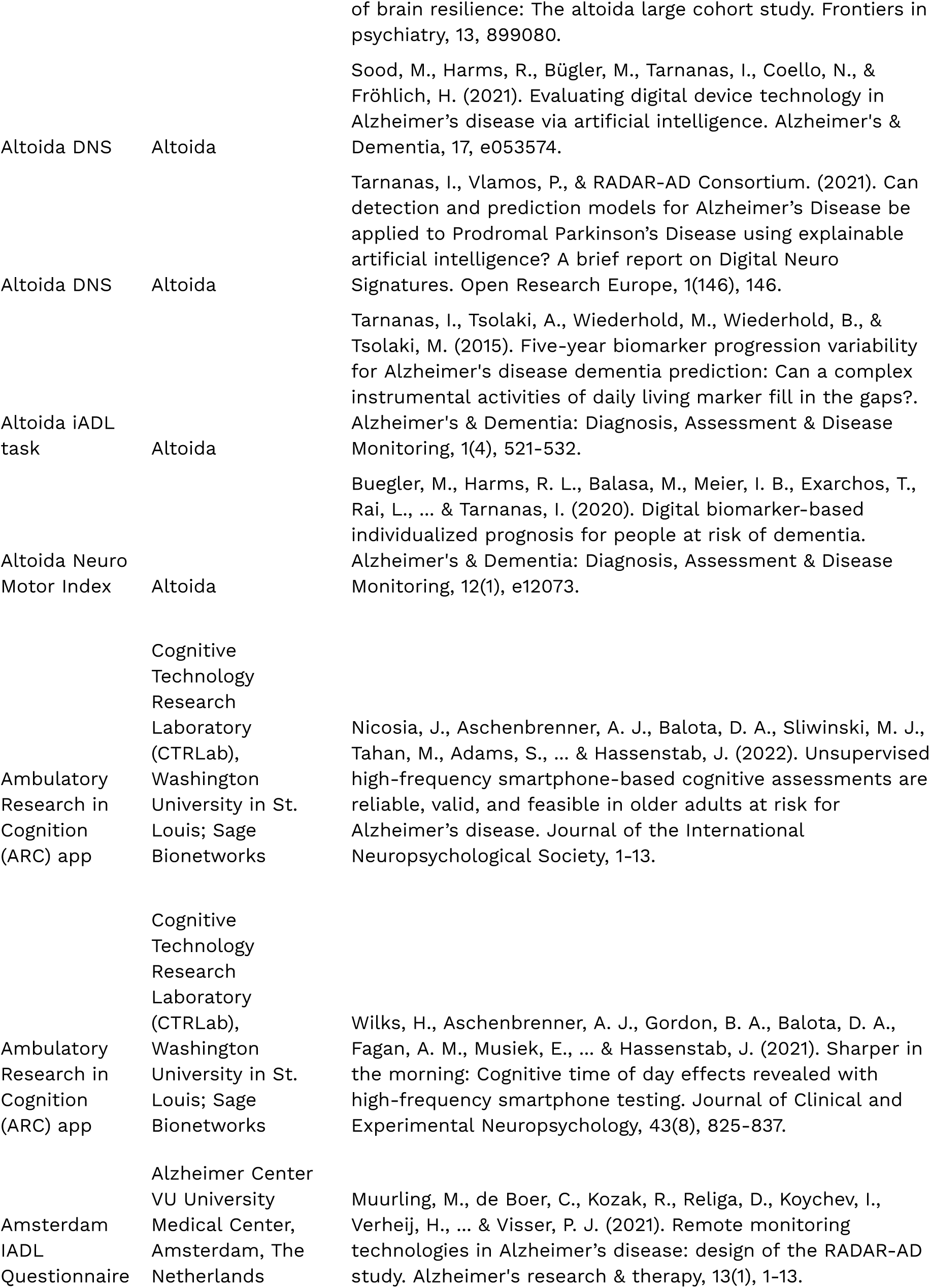
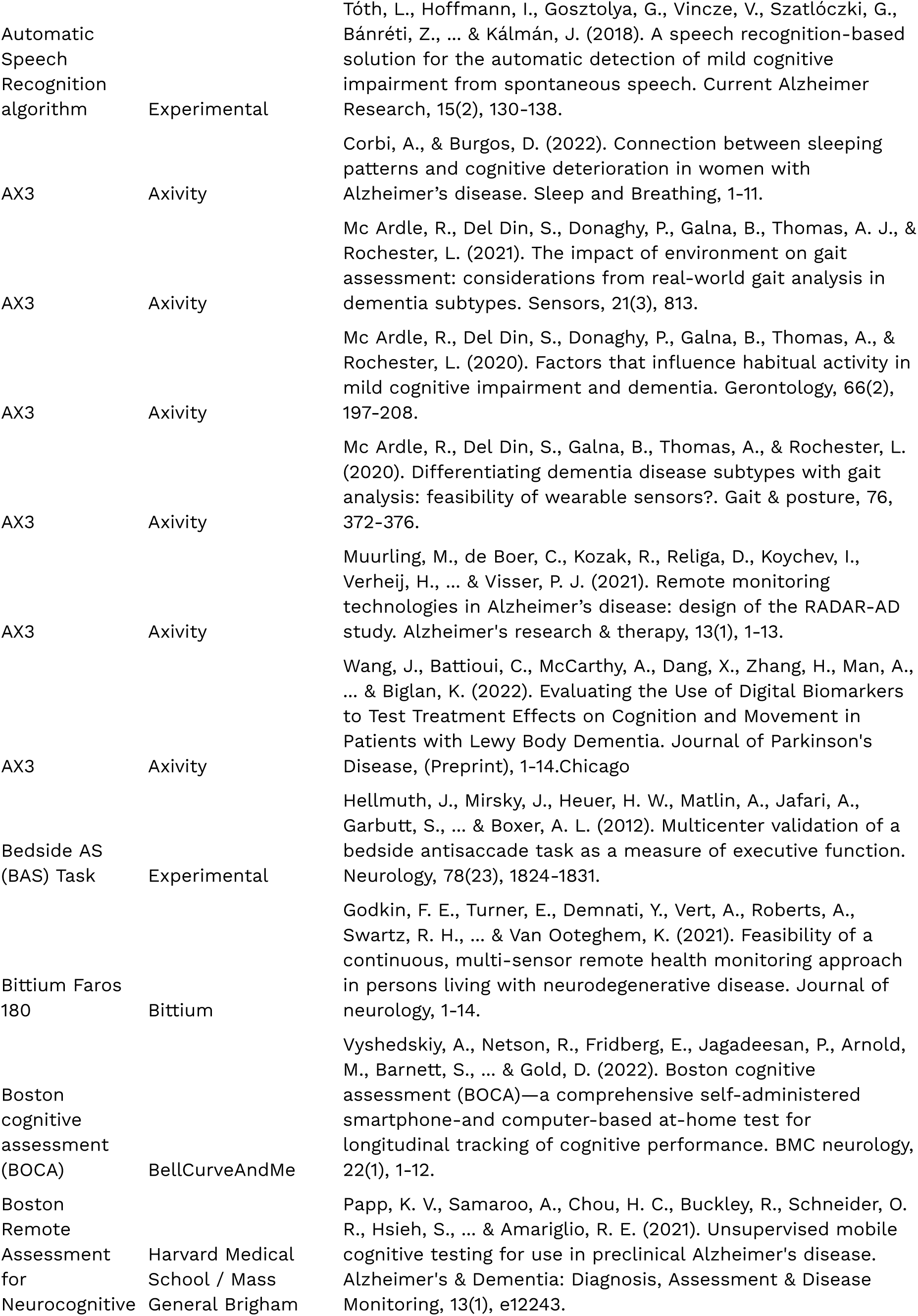
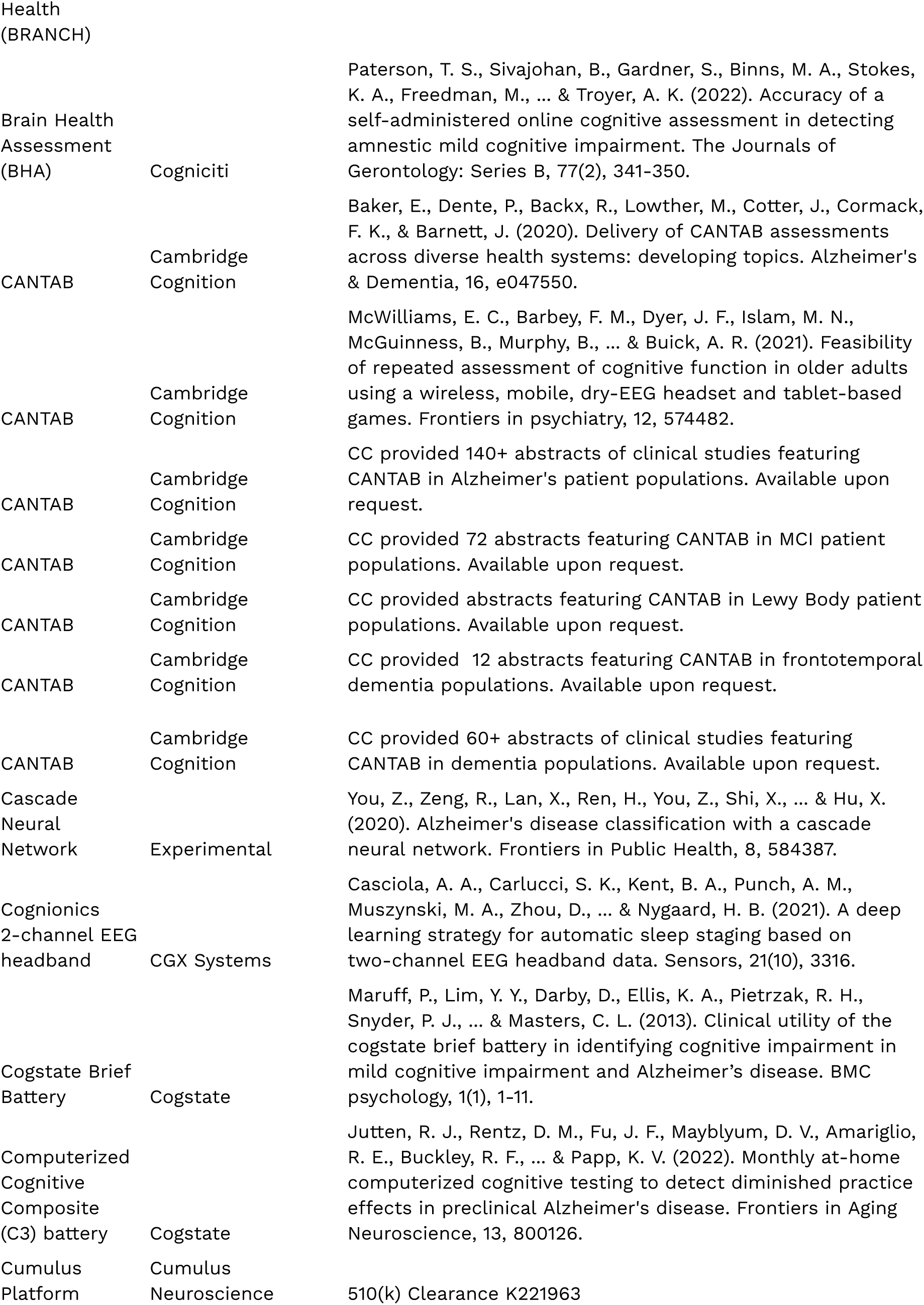
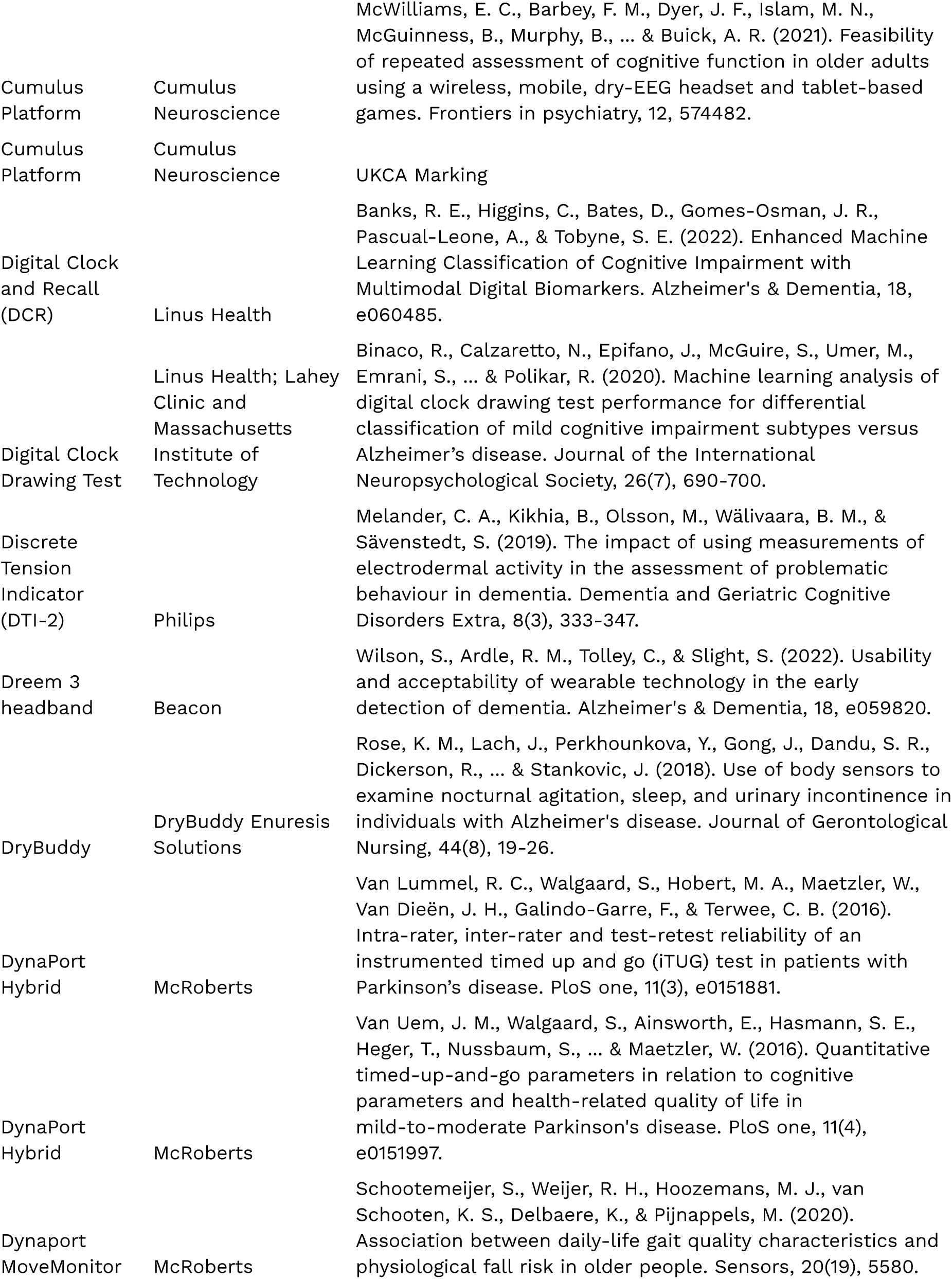
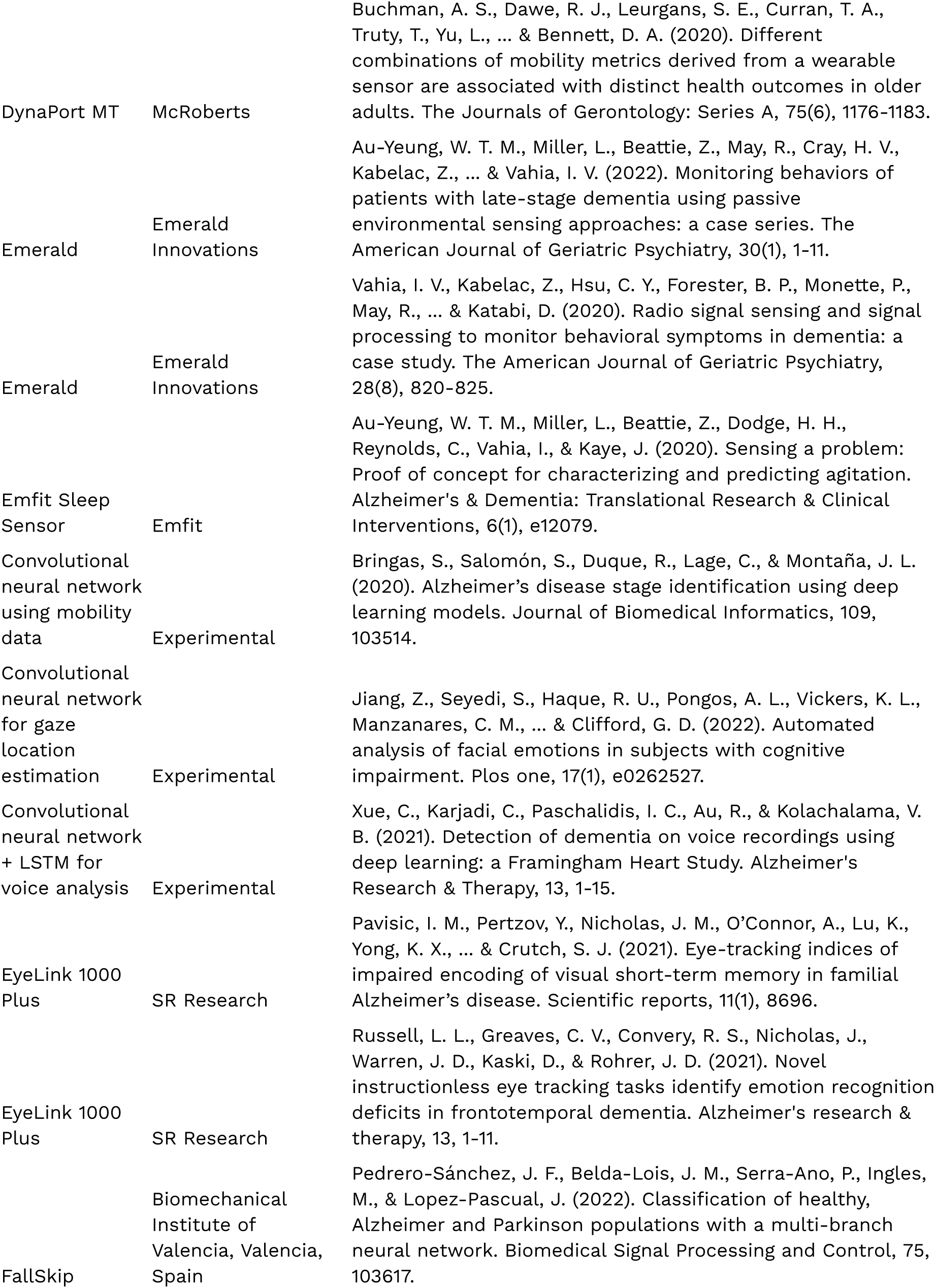
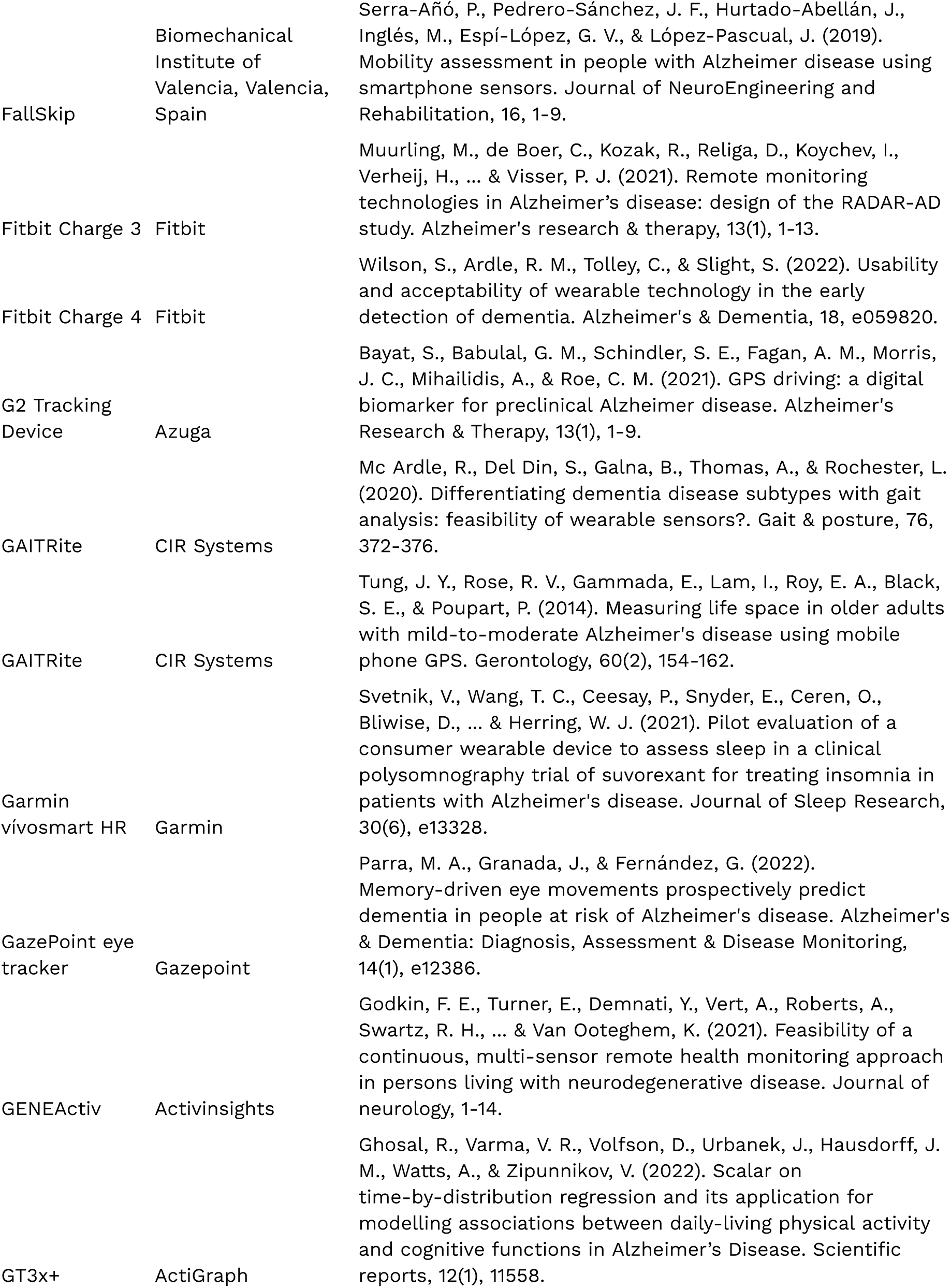
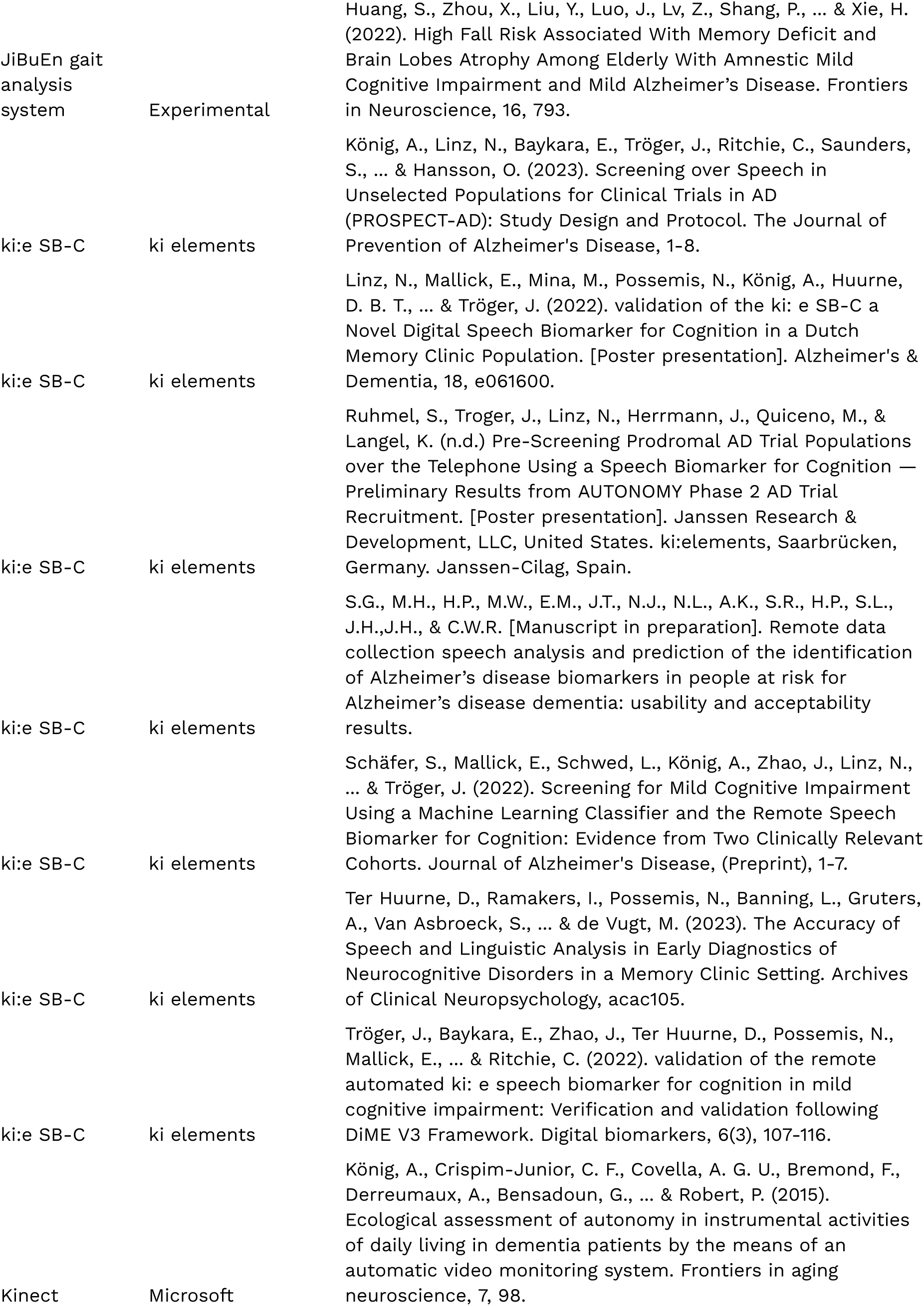
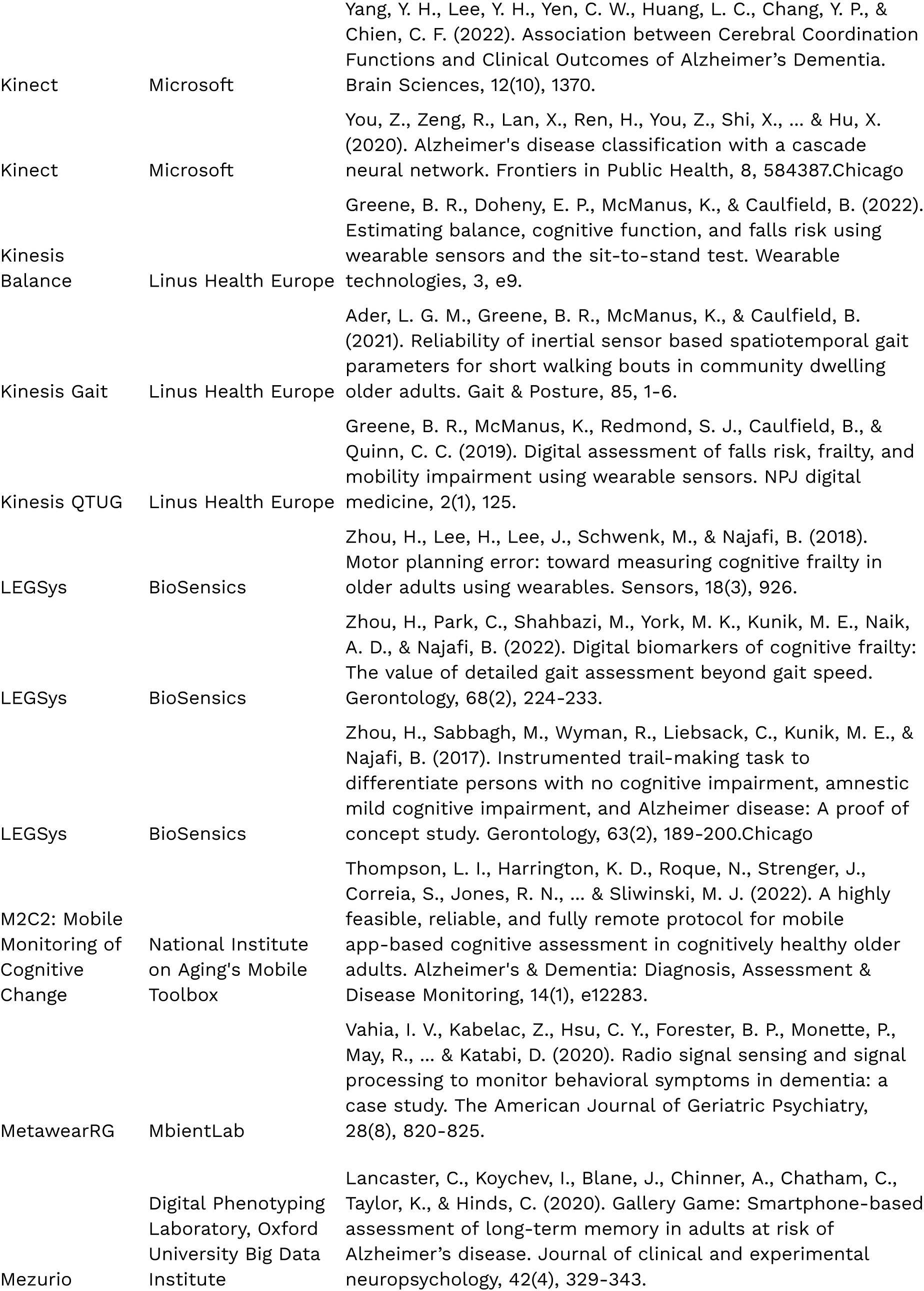
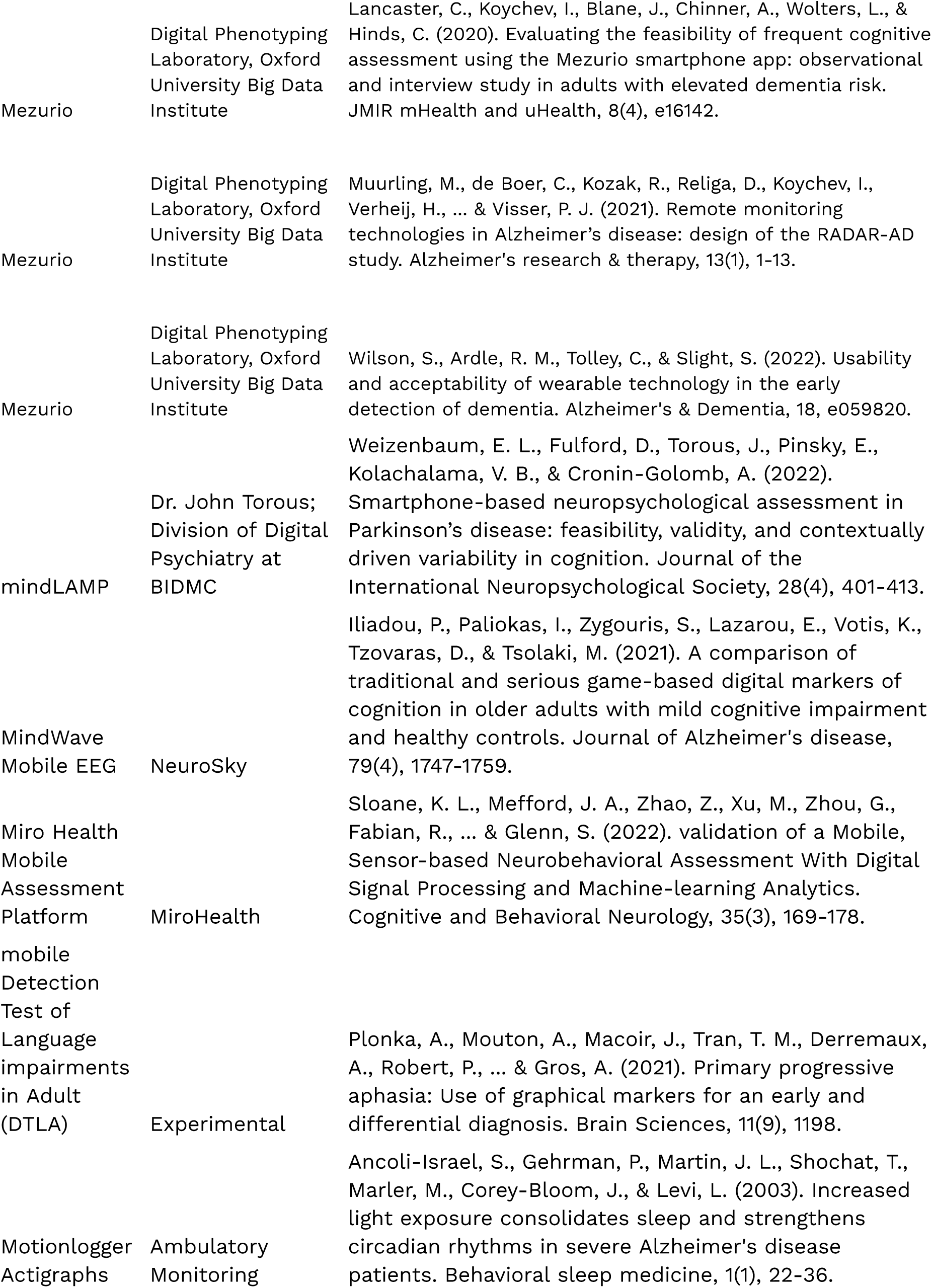
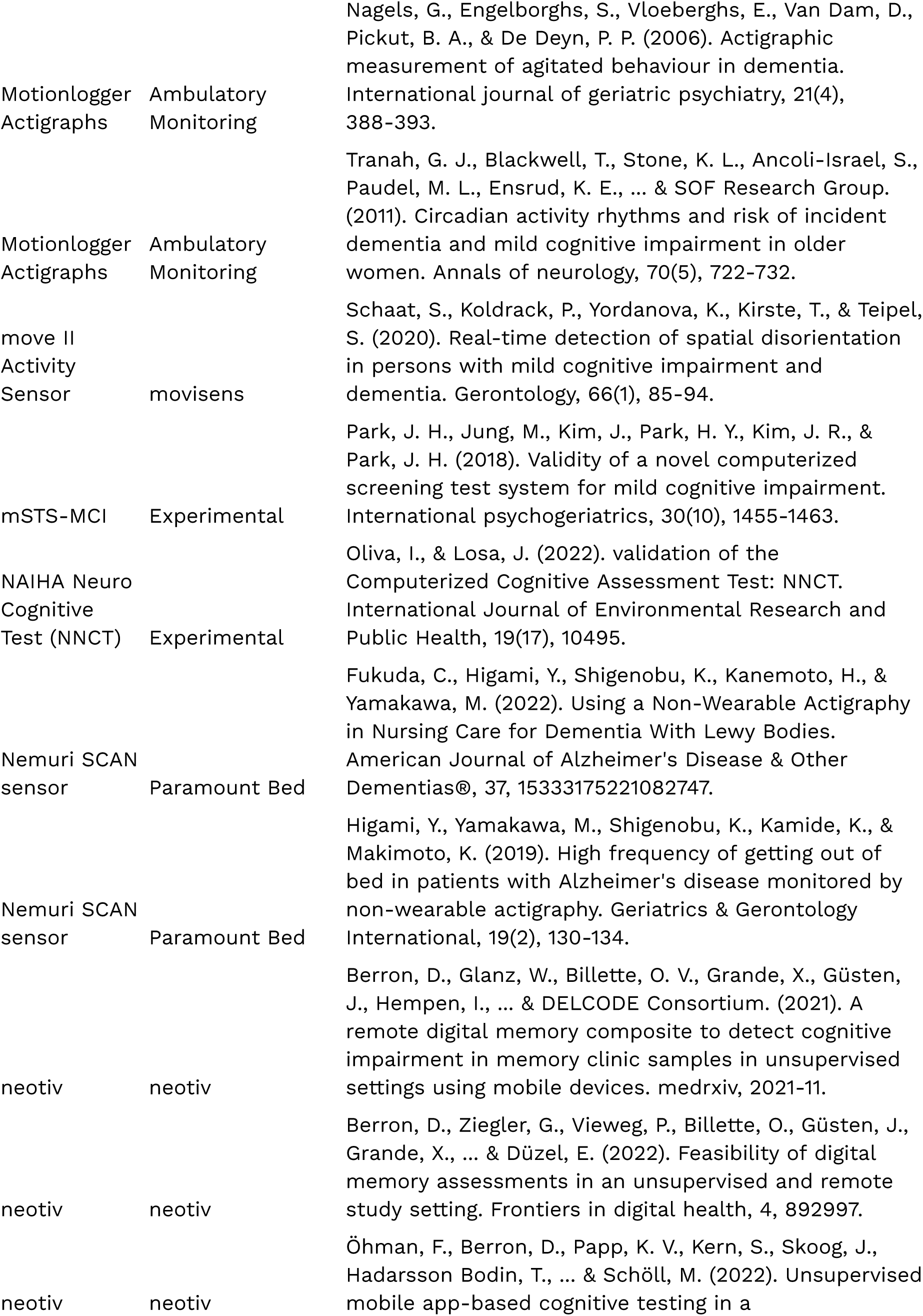
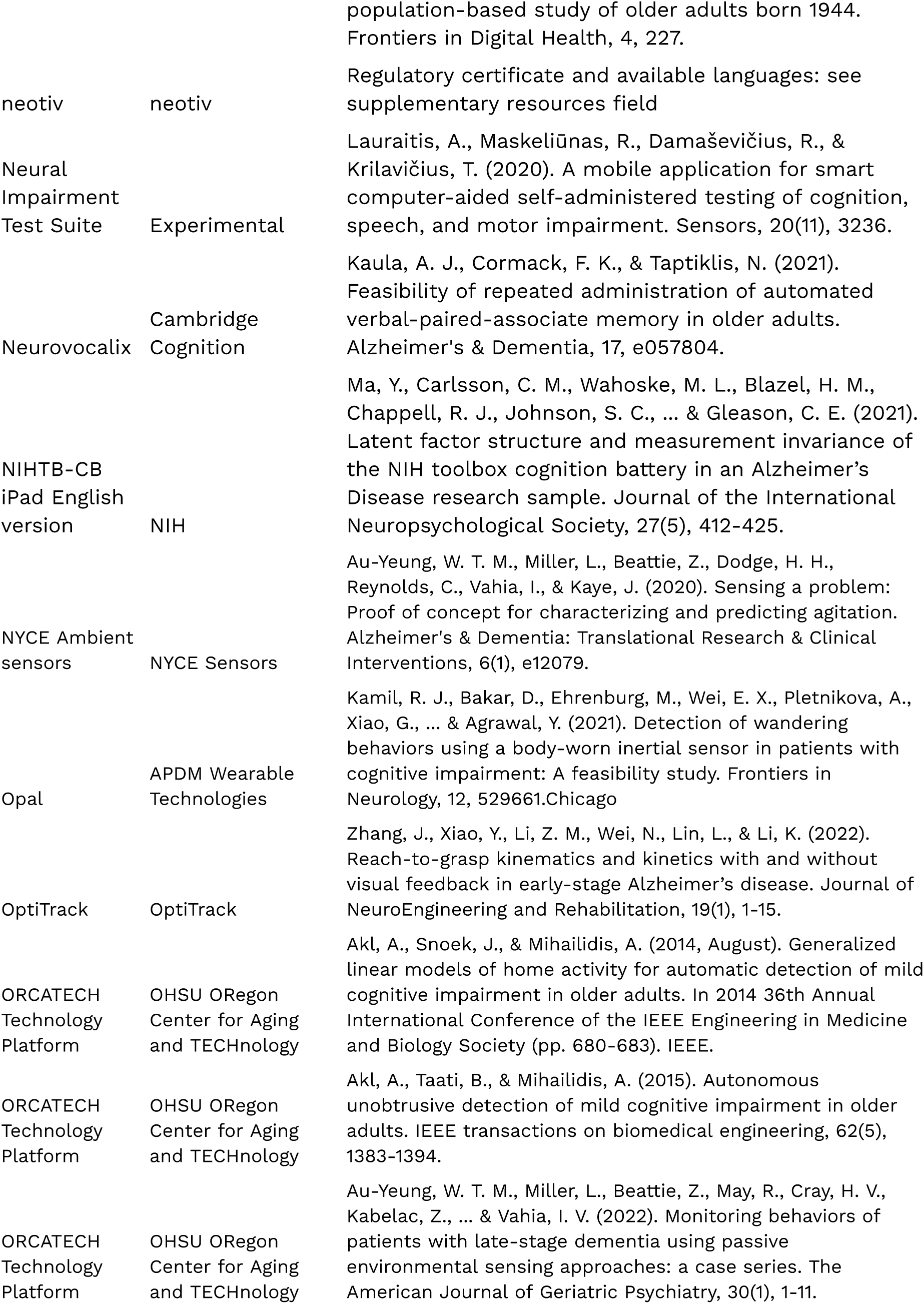
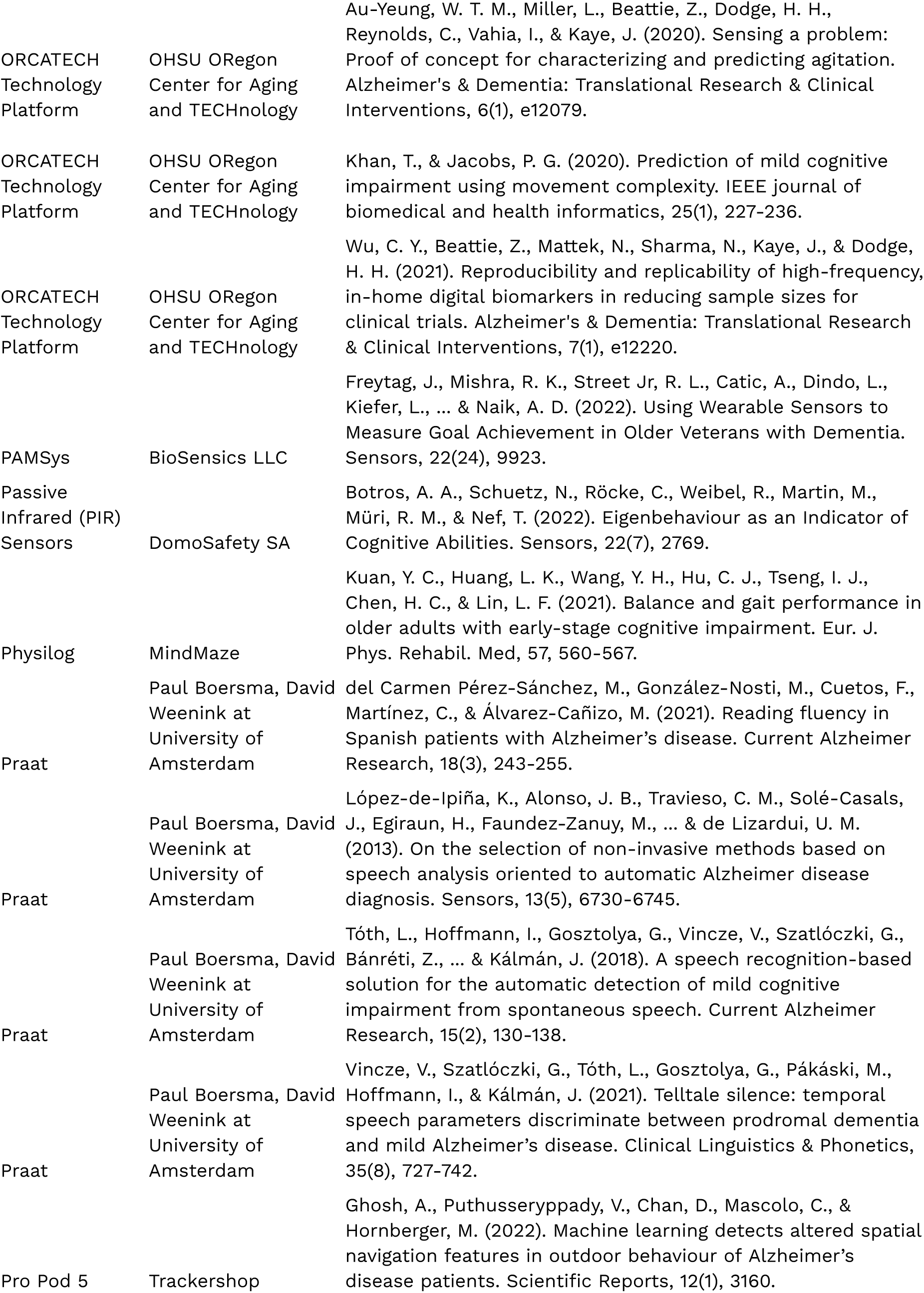
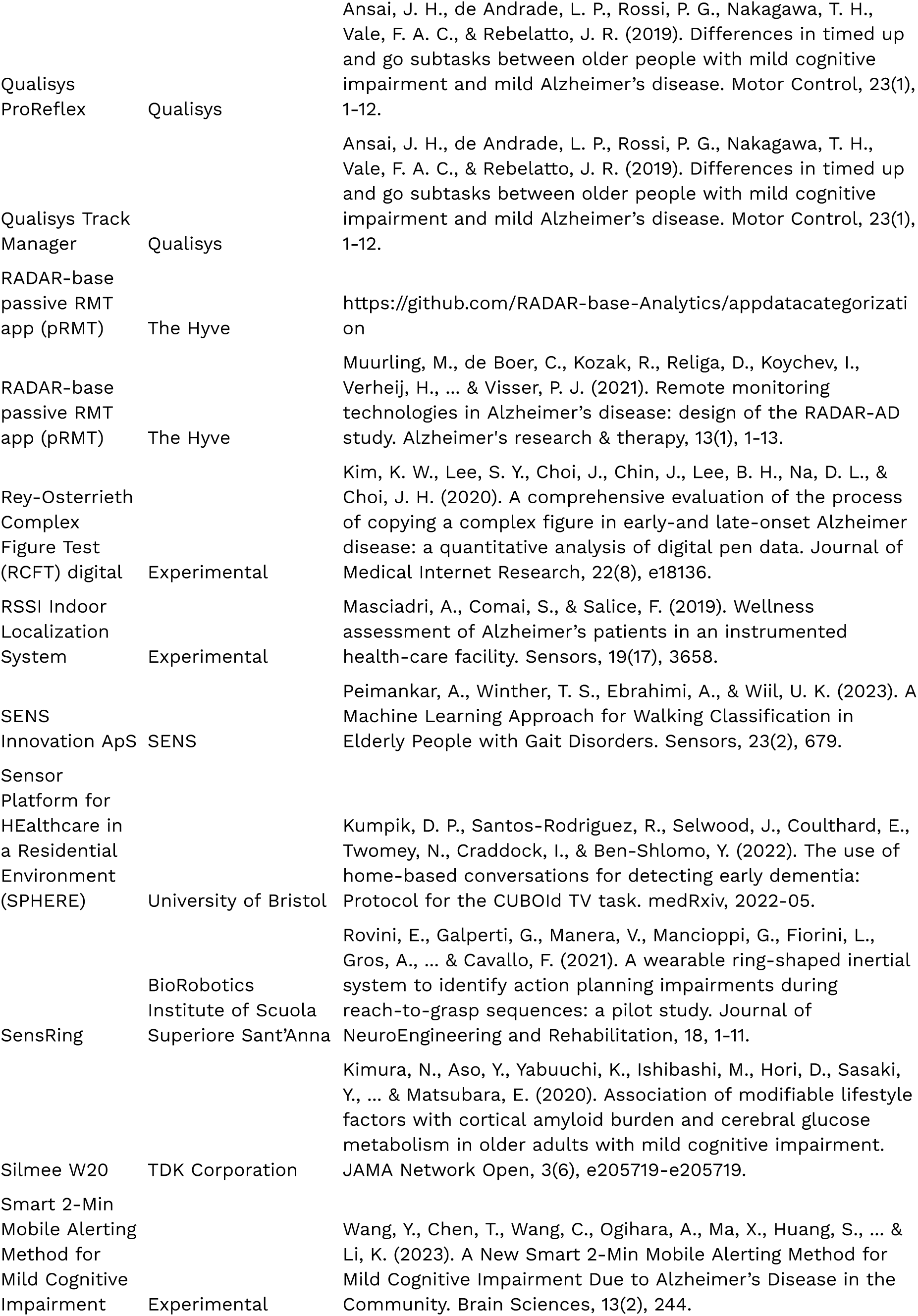
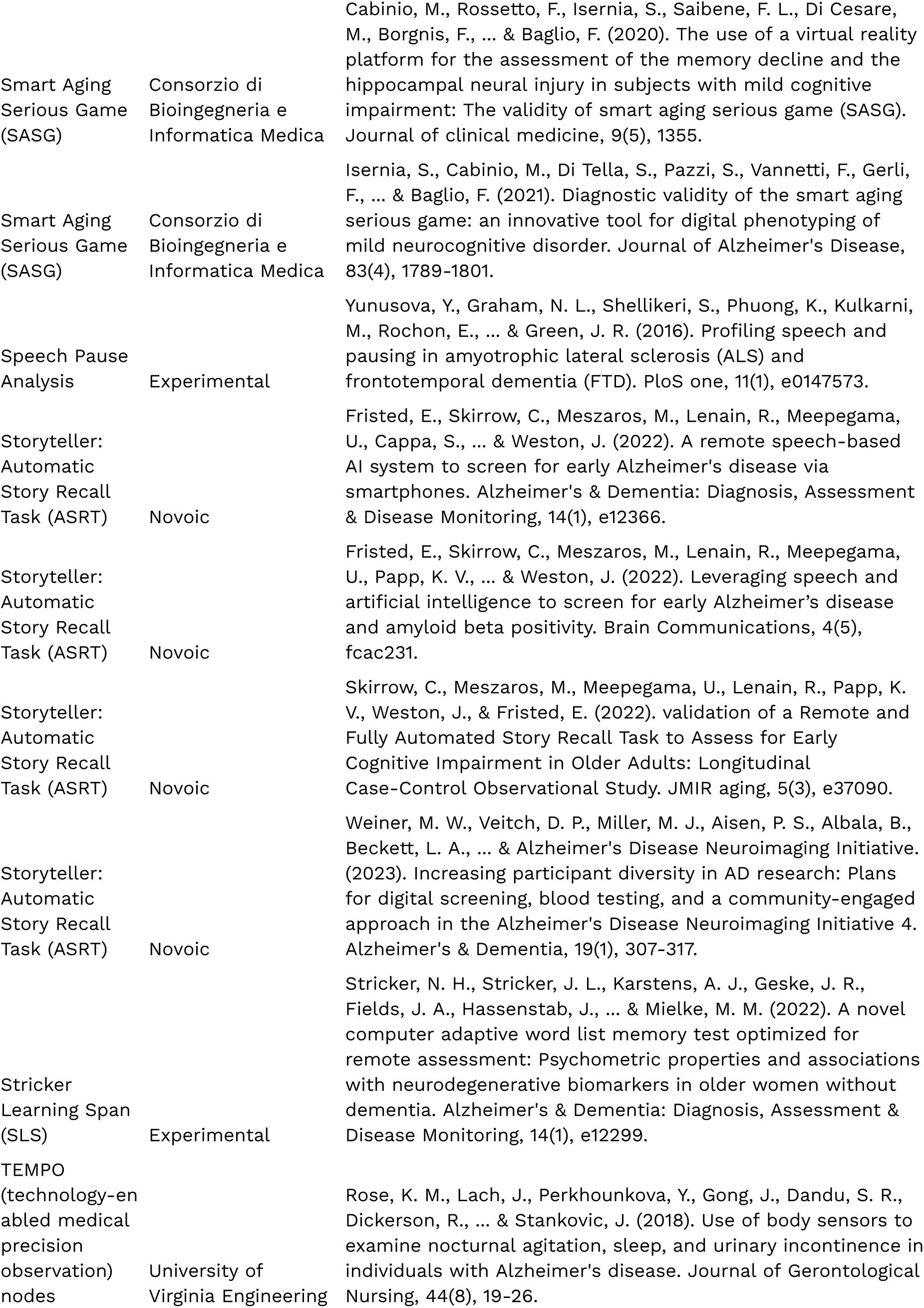
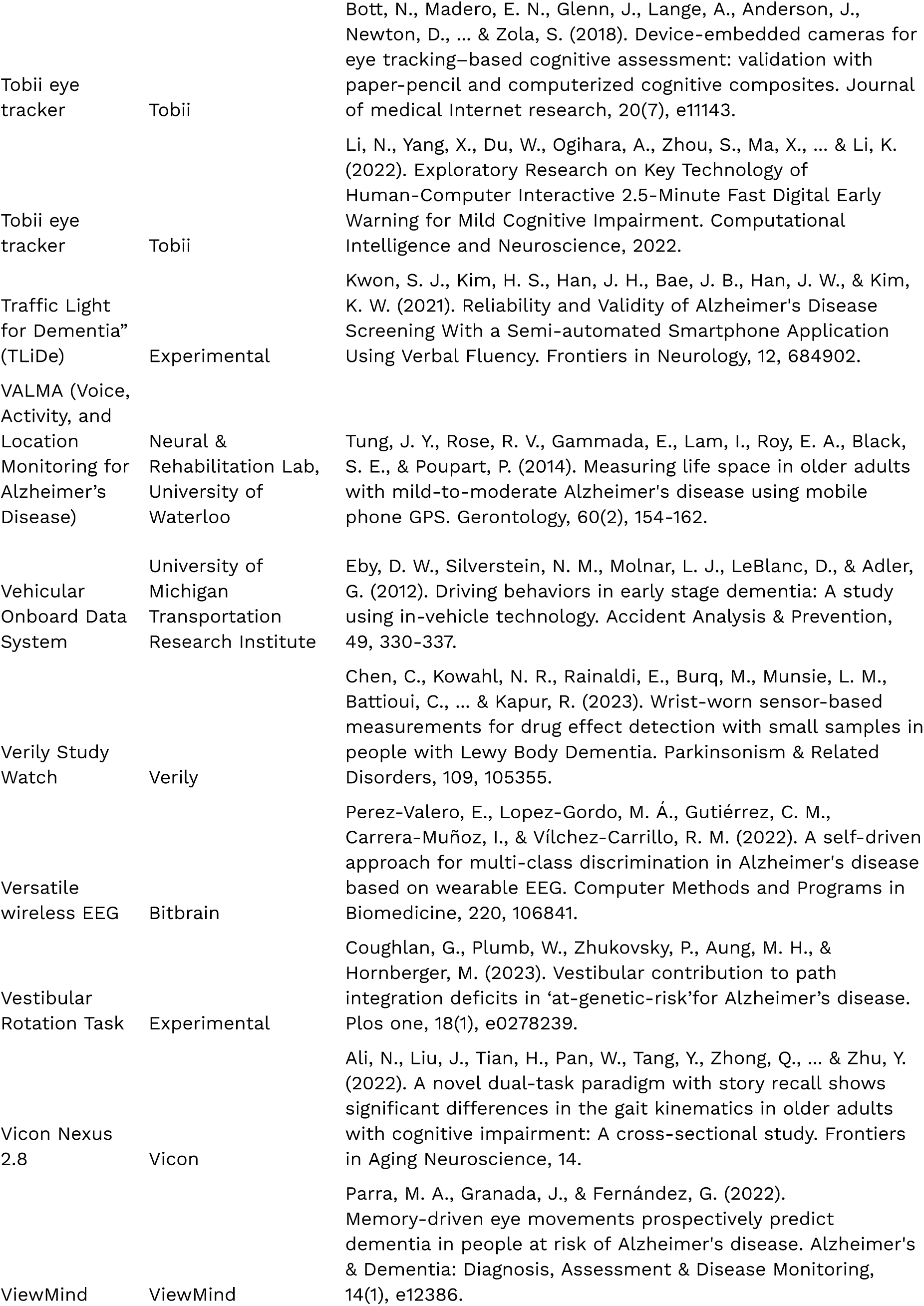
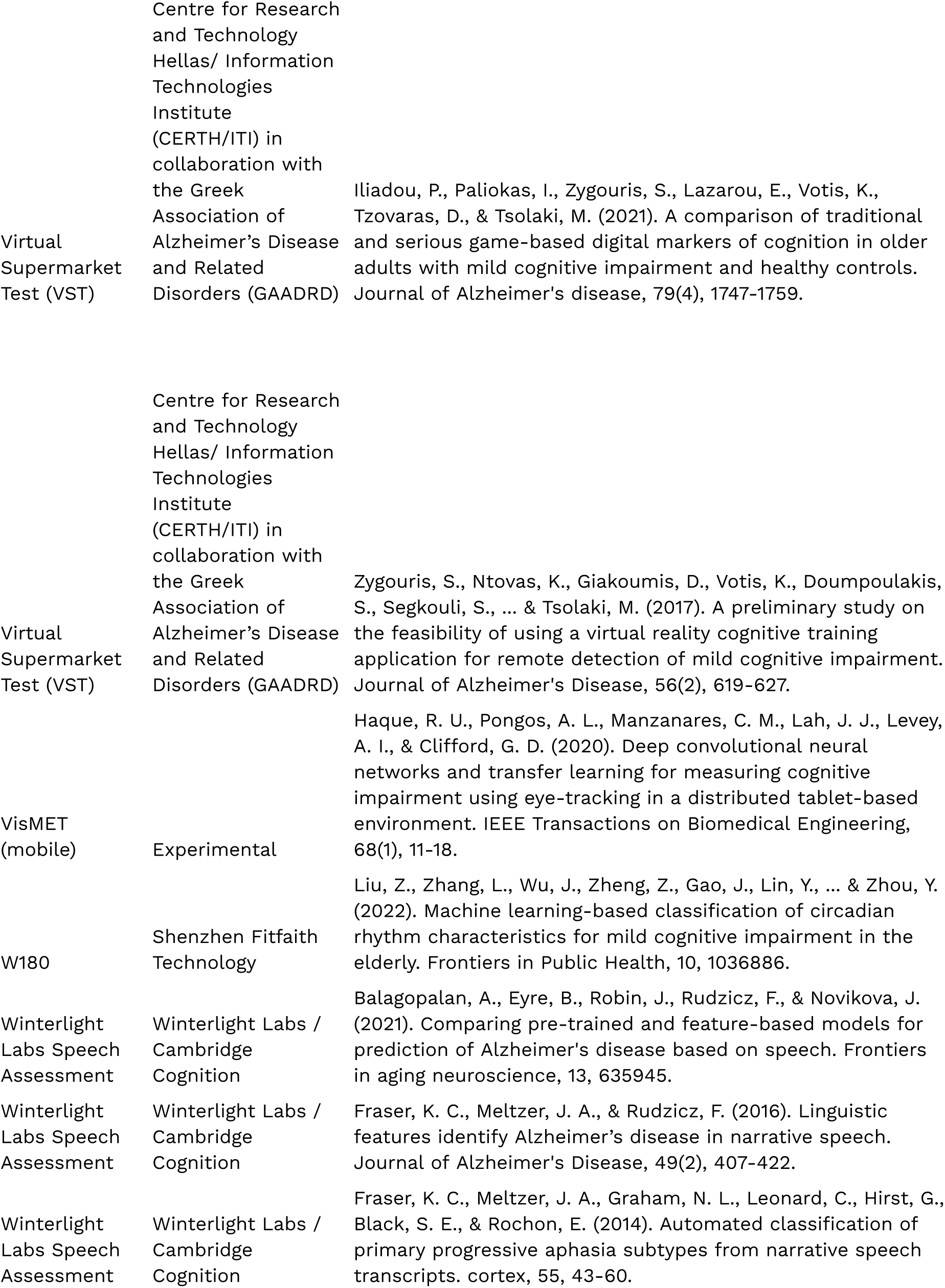
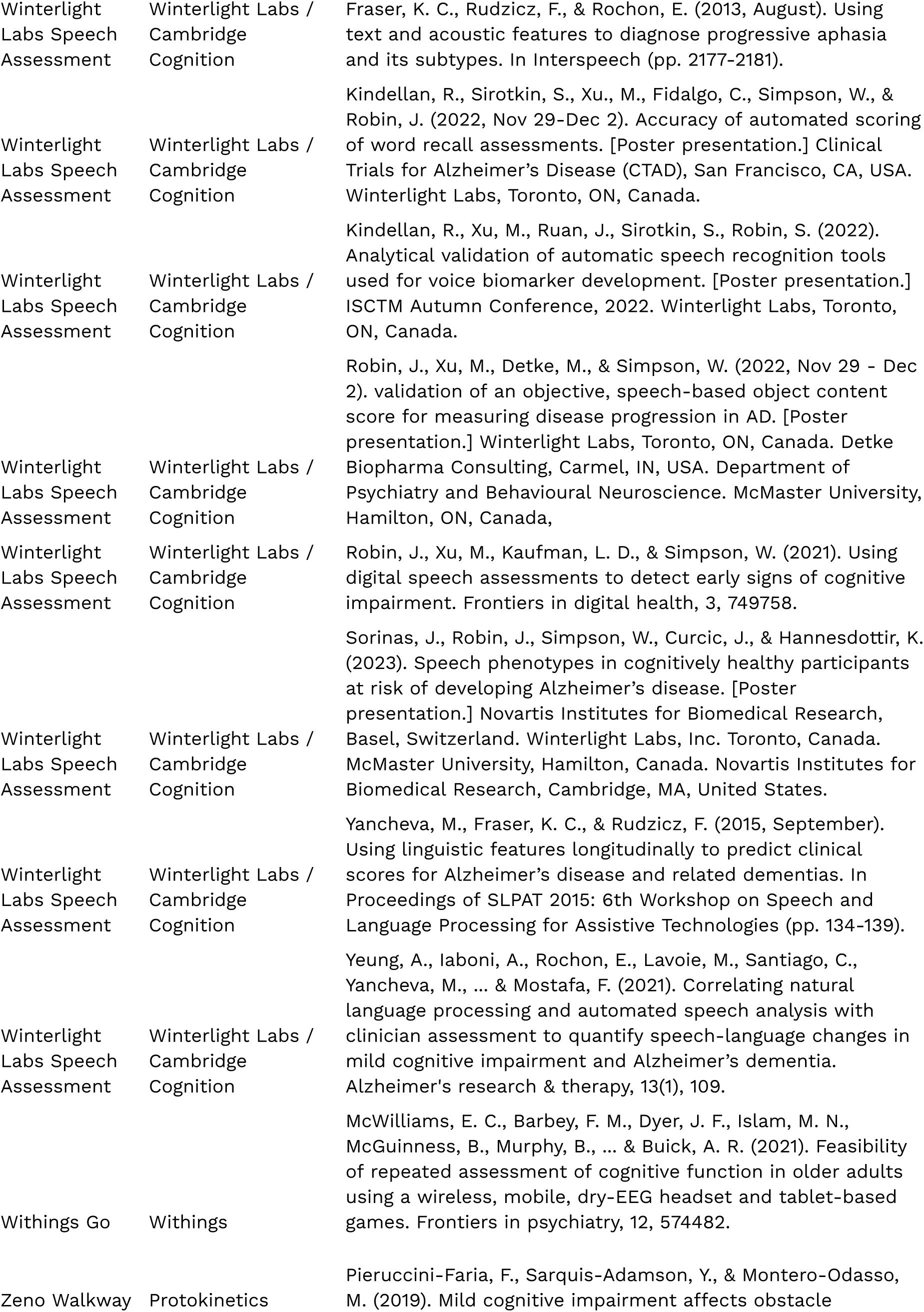
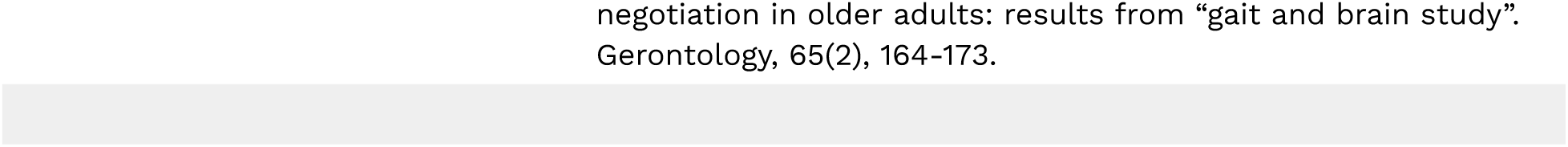

## Abbreviations

AD: Alzheimer’s disease
ADRD: Alzheimer’s disease and related dementias
DATAcc: Digital Health Measurement Collaborative Community
DHT: Digital health technology
EVIDENCE: <u>EV</u>aluatIng connecte<u>D</u> s<u>EN</u>sor te<u>C</u>hnologi<u>E</u>s
V3: Verification, analytical validation, and clinical validation

## Acknowledgments

We appreciate the support of our collaborating partners and experts who comprise the DATAcc by DiMe ADRD Project working group and supported this manuscript: Abbvie; Altoida, Alzheimer’s Drug Discovery Foundation, Aural Analytics, Biogen, BioSensics, Boston University, Cambridge Cognition, Cogniant Pte Ltd, Cognito Therapeutics, Cogstate, Cumulus Neuroscience, Eisai, Eli Lilly & Company, Gates Ventures, ki:elements, Koneksa, Linus Health, Luca Health, Medidata, Merck, Northwestern University, Oregon Health & Science University, Roche, Sage Bionetworks, VivoSense, Winterlight Labs.

## Author Contributions

All authors contributed to the conceptual development of this manuscript. All authors have approved the final submitted version and agree to be accountable for all aspects of the work presented here.

## Graphics

**Table 1.**
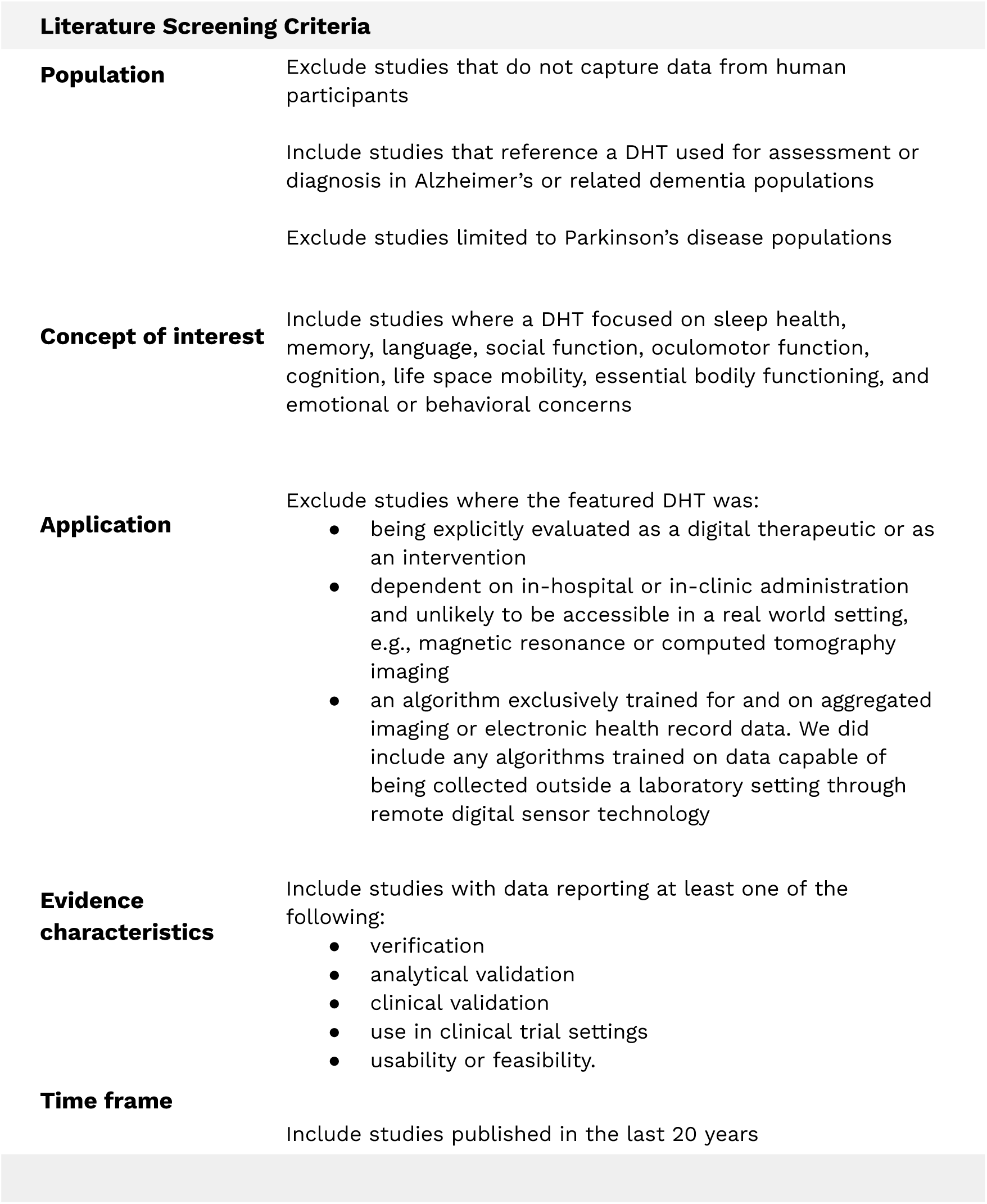
Inclusion/exclusion criteria applied to identified papers in the landscape review.

**Table 2.**
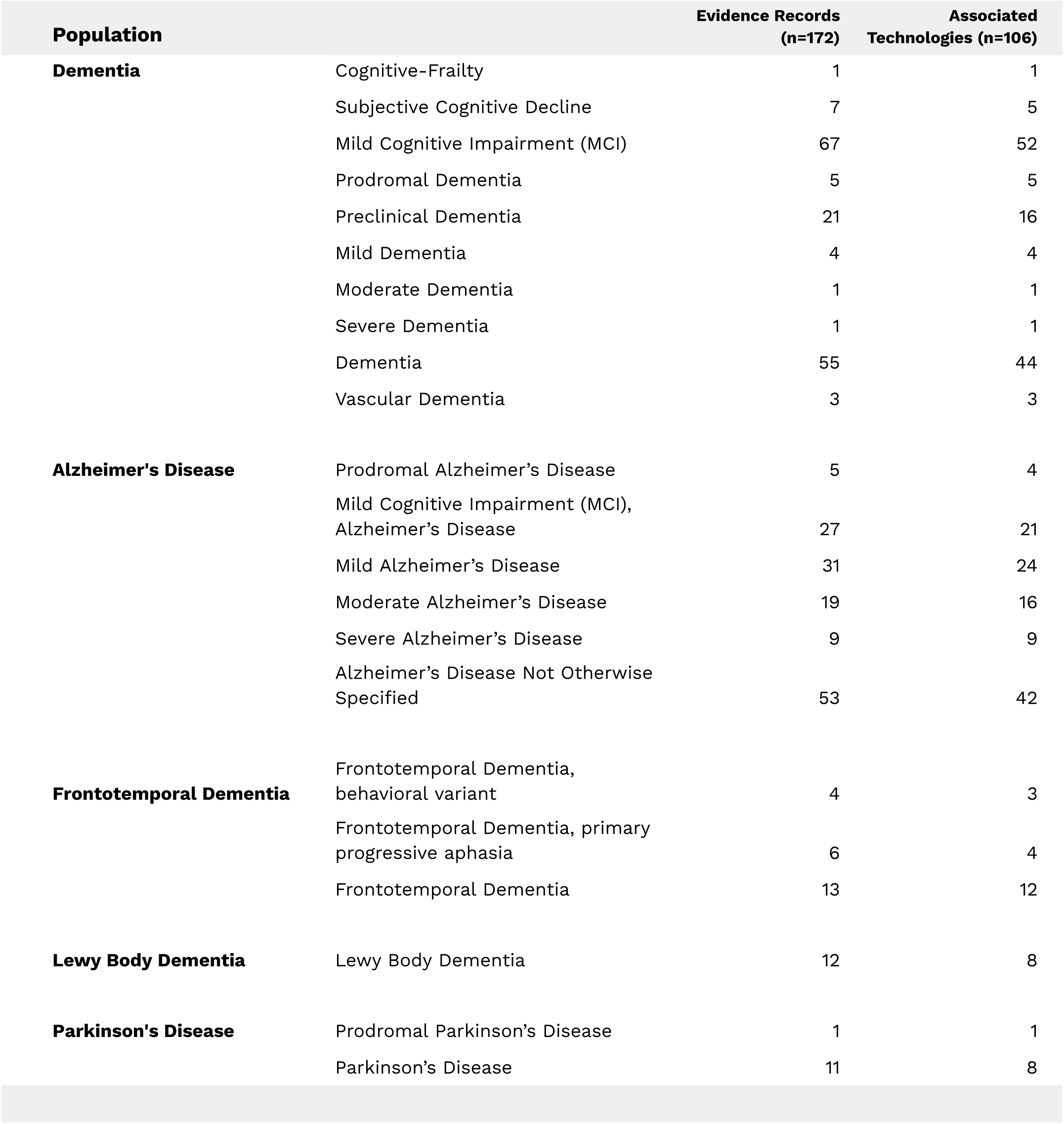
Frequencies of population areas of focus in the library, with descriptors as reported in the literature. Many pieces of evidence reference multiple diagnostic classifications.

**Table 3.**
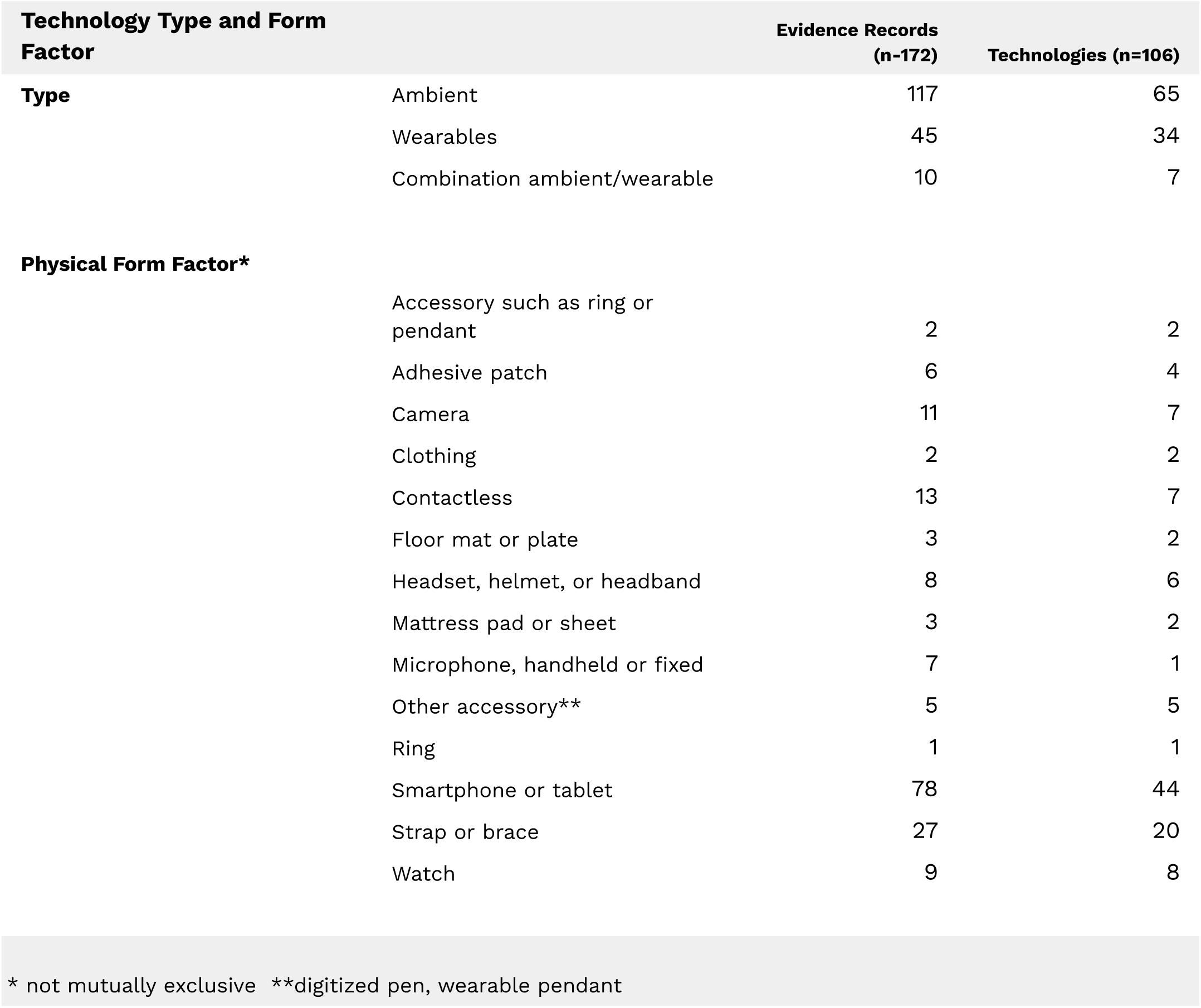
Prevalent technology types and form factors represented in the ADRD DHT library. Ambient includes environmental and software or application-based technologies.

**Table 4.**
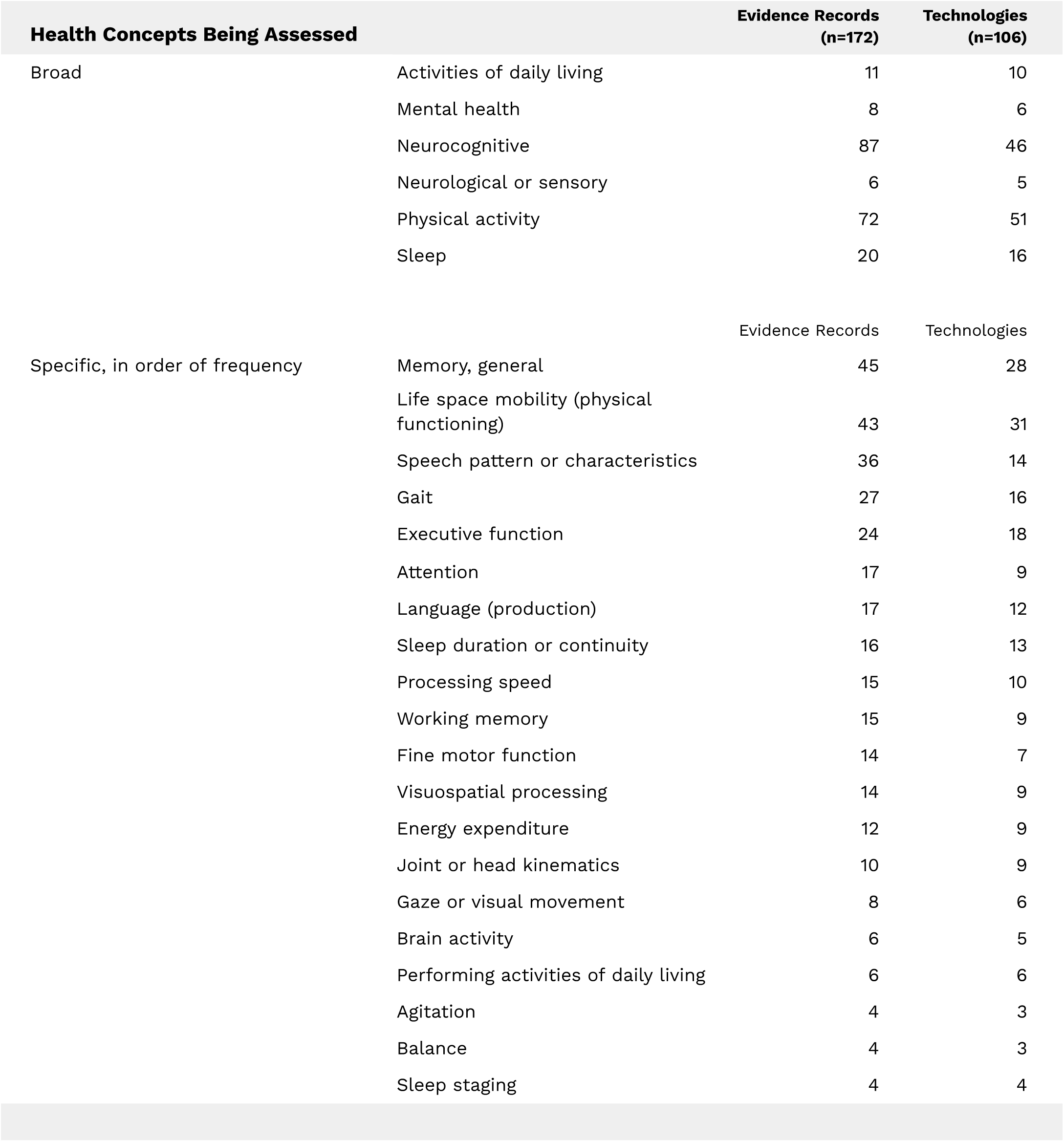
Prevalent concepts of interest in the ADRD DHT library. These are not mutually exclusive. These figures represent frequency in library evidence along with the number of corresponding technologies as of October 2023.

